# Development and Validation of an Asthma Policy Model for Canada: Lifetime Exposures and Asthma outcomes Projection (LEAP)

**DOI:** 10.1101/2024.03.11.24304122

**Authors:** Tae Yoon Lee, John Petkau, Kate M. Johnson, Stuart E. Turvey, Amin Adibi, Padmaja Subbarao, Mohsen Sadatsafavi

## Abstract

**Purpose:** To develop Lifetime Exposures and Asthma outcomes Projection (LEAP), a reference policy model for evaluating health outcomes and costs of asthma interventions and policies for the Canadian population.

**Methods:** Following the best practice guidelines for development, we first created a conceptual map with a steering committee of clinician experts and economic modelers through a modified Delphi-process. Following the committee’s recommendations and given the multidimensionality of risk factors and the need for modeling realistic aspects (e.g., gradual market penetration) of adopting health technologies, we opted for an open-population microsimulation design. For the first version of the model, we concentrated on several key risk factors (age, sex, family history of asthma at birth, and exposure to antibiotics in the first year of life) from the concept map.

The model consists of five intertwined modules: 1) demographic, 2) risk factors, 3) asthma occurrence, 4) asthma outcomes, and 5) payoffs. The demographic module, including birth, mortality, immigration, and emigration, was based on sex– and age-specific estimates and projections from Statistics Canada. The distributions of risk factors, including family history of asthma and exposure to antibiotics, were estimated from population-based administrative databases and a population-based longitudinal birth cohort. To estimate parameters in the asthma occurrence (prevalence, incidence, reassessment) and asthma outcomes (severity, symptom control, exacerbations) modules, we performed quantitative evidence synthesis. Costs and utility weights were obtained from the literature. We conducted multiple face and internal validation assessments.

**Results:** LEAP is capable of modeling asthma-related health outcomes at the individual and aggregate levels from 2001 onwards. Face validity was confirmed by checking the structure, equations, codes, and results. We calibrated and internally validated the age-sex stratified demographic projections to the estimates and projections from Statistics Canada, the age-sex stratified asthma prevalence to the administrative data, and the asthma control levels and exacerbation rates to the estimates from the literature.

**Conclusions:** LEAP is the first reference Canadian asthma policy model that emerged from identified needs for health policy planning for early interventions in asthma. As an open-source and open-access platform, LEAP can provide a unified framework under which different interventions and policies can be consistently compared to identify those with the highest value proposition.

**Funding:** This study was funded by a research grant from the Canadian Institutes of Health Research and Genome Canada (274CHI). The funders had no role in any aspect of this study and were not aware of the results.

**Ethics:** This study was approved by the institutional review board of the University of British Columbia, Vancouver (H22-00571).

## Background

Asthma is an early-onset, chronic disease of the airways affecting over 3 million Canadians and 339 million people worldwide (GBD 2019 Diseases and Injuries Collaborators, 2020; Public Agency of Canada, 2018; Soriano et al., 2020). It is one of the leading causes of emergency department visits, hospital admissions, missed school days for children, and loss of work productivity for adults. It greatly reduces the quality of life of patients and their families (Bahadori et al., 2009; Ehteshami-Afshar et al., 2016; FitzGerald et al., 2020). By all accounts, asthma imposes a substantial economic and humanistic burden to individuals with asthma and society.

Due to the absence of a cure for asthma, the contemporary management of asthma is focused on achieving and maintaining asthma control (Global Initiative for Asthma, 2023; Lommatzsch et al., 2023). However, emerging evidence promises the development of innovative interventions that might reduce the risk of asthma onset in children. For example, researchers have found an association between the risk of asthma and exposure to antibiotics during infancy, a relationship that seems to be mediated by the gut microbiome (Hoskinson et al., 2023; Patrick et al., 2020). This finding corroborates the microflora hypothesis (Stiemsma and Turvey, 2017) which states that exposure to certain gut microbes in early life is critical for developing a robust immune system. Thus, interventions and policies, such as reducing unnecessary exposure to antibiotics, may prevent the development of asthma among children.

Efficient health policy-making relies on projections of future consequences of decisions. Hence, epidemiological forecasting, burden of disease projections, and economic evaluations are foundational to evidence-informed policymaking. Of particular importance in health policymaking is economic evaluation of competing interventions. This is because the implementation of any new intervention or policy is associated with ‘opportunity costs.’ In this context, the opportunity cost of implementing a particular intervention is the health benefits that could have been obtained if the resources had been used to implement another intervention. Stakeholders (e.g., the government and patients) who have limited resources must know the ‘value for money’ potential of these competing interventions to make informed decisions.

For the most part, projecting health outcomes at the population level and conducting economic evaluations require creating a decision-analytic framework of the disease of interest and quantifying the impact of interventions under evaluation (Buxton et al., 1997). Such a framework is often realized as a computer simulation model that allows one to: 1) explicitly describe the complex interplay of risk factors, interventions, and health and economic outcomes, 2) synthesize data from multiple sources (e.g., effectiveness from clinical trials and evidence on health services use from electronic medical records) in a unified framework, 3) project long-term, policy-relevant metrics based on intermediate clinical outcomes or outcomes from studies with shorter follow-up times, and 4) project the outcomes under different ‘what-if’ scenarios (e.g., how many asthma cases will be averted in the next 20 years if a national antibiotic stewardship program is implemented).

A recent scoping review and an earlier systematic review concluded that current decision-analytic asthma models generally do not consider the multifaceted and heterogeneous nature of the disease, lack transparency, lack sufficient granularity to model the nuances of interventions (e.g., imperfect adherence to treatments), and do not fully adhere to recommendations on modeling and outcomes (Adibi et al., 2021; Ehteshami-Afshar et al., 2019). Lack of high-quality economic evidence is likely to hinder the implementation of interventions that could substantially reduce the burden of asthma. Customarily, a new computer model is developed to conduct policy analysis and economic evaluation for each set of new health interventions. In recent years, this approach has been criticized for several reasons (Afzali et al., 2013). Such ‘piecemeal’ modeling is deemed inefficient, because different computer models are sometimes developed and used for the same disease, resulting in a loss of analytic resources. Additionally, such *de novo* models are prone to be inconsistent in terms of model structure, evidence, and underlying assumptions. There is an increasing recognition that de novo models often lack transparency, making them inaccessible to other users (Kent et al., 2019). An alternative approach to address these issues is the use of a ‘reference model’ that serves as a unified framework for evaluating different interventions for the same disease (Afzali et al., 2013). The reference model must be transparent enough so that users can understand its model structure and assumptions and use it with confidence. The need for a national reference policy model for asthma emerged from discussions among key stakeholders in Canada (Legacy for Airway Health, 2020). To address this knowledge gap, our overarching goal was to develop and validate a reference policy model for evaluating interventions for asthma in Canada.

## Methods

Our model development and validation underwent the following stages, in accordance with the best standards set forth by the Professional Society for Health Economics and Outcomes Research (ISPOR) – Society of Medical Decision Making (SMDM) Modeling Good Research Practices Task Force (Briggs et al., 2012; Caro et al., 2012; Eddy et al., 2012; Karnon et al., 2012). First, we convened a group of methodologists and clinical experts to create a conceptual map. Based on the conceptual map, we selected an appropriate model structure and identified key model components. We performed several analyses to generate and synthesize evidence required for the model and calibrated the model to the Canadian population. We then implemented the model as open-source, open-access software. Lastly, we carried out validation studies to test the validity of model assumptions and implementation.

As recommended, the process started with a concept map, developed by deep engagement with a steering committee of economic modelers, allergists, and respirologists across Canada (Figure 1). At the time, interventions of interest were early preventive strategies. As such, the group’s focus was on childhood asthma. The model concept, which emerged through a modified Delphi process (details are provided in Adibi et al. (2021)), includes three major groups of risk factors (including unmodifiable ones) related to diagnosis of childhood asthma: patient characteristics, family history, and environmental factors.

**Figure 1:**
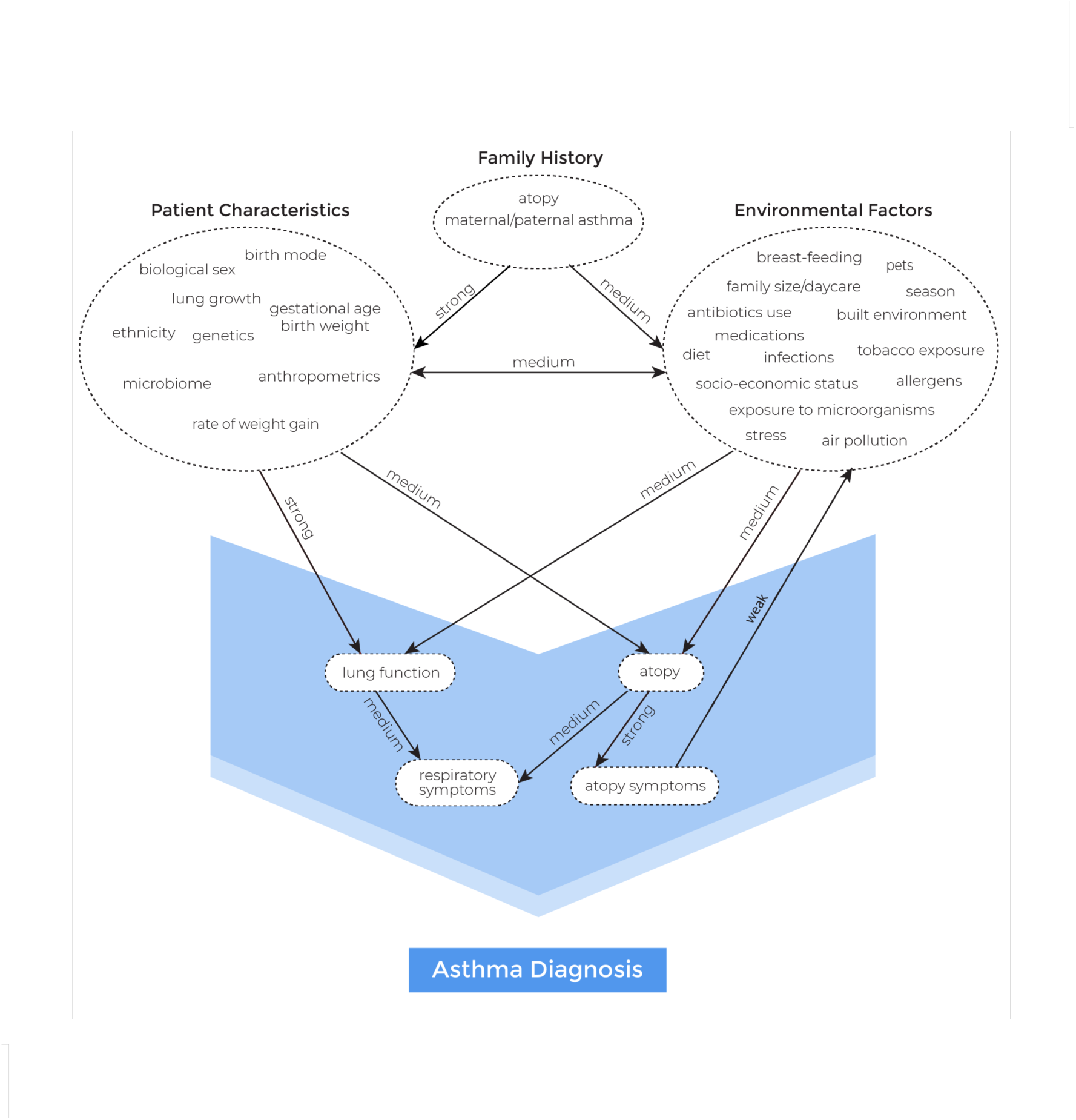
Concept map of childhood asthma. An arrow indicates the direction of the relationship, along with the strength written above it (used with permission from BMJ).

Our implementation of this conceptual model is as an open-population discrete-time microsimulation, in alignment with recommendations from the steering committee (Adibi et al., 2021). In a microsimulation model, a virtual individual is created to represent a person in the population of interest. In each time cycle, actions and behaviors of the individual are simulated based on pre-defined rules, and the attributes and disease characteristics of the individual are updated accordingly. The microsimulation model is capable of accommodating the multidimensionality of risk factors and a high number of disease states in their transition probabilities. A virtual individual in our model represents a Canadian. We did not allow interactions between virtual individuals (hence not modeling vertical relationships in the model) and set the time cycle unit to annual as we concluded the annual time cycle strikes a right balance between granularity in simulation events over time and the computational demands of the planned analyses.

The model is open-population as it simulates the entire population over time, including population growth, aging, immigration, and emigration. This feature was deemed necessary to satisfy the need for modeling realistic aspects (e.g., gradual market penetration) of adopting health interventions for an immigrant welcoming country, such as Canada (Willekens and Van Imhoff, 2015). Correspondingly, the time horizon (i.e., the period over which the model can be run) is defined in calendar time. The maximum range of the time horizon is from 2001 (the earliest year for which we had access to data) to 2065 (the latest year for which population projections are available), enabling both retrospective (e.g., the impact of previous health interventions on the current burden of asthma) and prospective modeling. If the base year is set to a time after 2019 (the latest year in which the data on population structure is available), the model internally will simulate the 2019 population towards the base year.

We modeled labeled, rather than true, asthma states due to lack of evidence on the natural history of asthma regarding true states. Current evidence on asthma is mostly based on labeled asthma states (physician-diagnosis or International Classification of Disease [ICD] codes). With better understanding of asthma and better technology that enables objective diagnostic testing, we envision obtaining and incorporating evidence on true asthma states in the future.

## Model components

The asthma model consists of five intertwined modules: 1) demographics, 2) risk factors, 3) asthma occurrence, 4) asthma outcomes, and 5) payoffs (costs and utilities). The core of the model can be conceptualized as a series of stochastic structural equations relating risk factors to each other and to events of interest (Figure 2). For the first version of the model, guided by consultation with the steering committee, we concentrated on several key risk factors from the concept map: age, sex, family history of asthma at birth, and infant (<1 year old) exposure to antibiotics. This represents at least one risk factor from each risk factor group (from Figure 1), and is aligned with the first identified use case for the model (i.e., the evaluation of infant antibiotic use on burden of asthma). Table 1 shows the summary of the structural equations by modules in the asthma model. Parameter values are provided in a separate table online (https://github.com/tyhlee/LEAP.jl).

**Figure 2:**
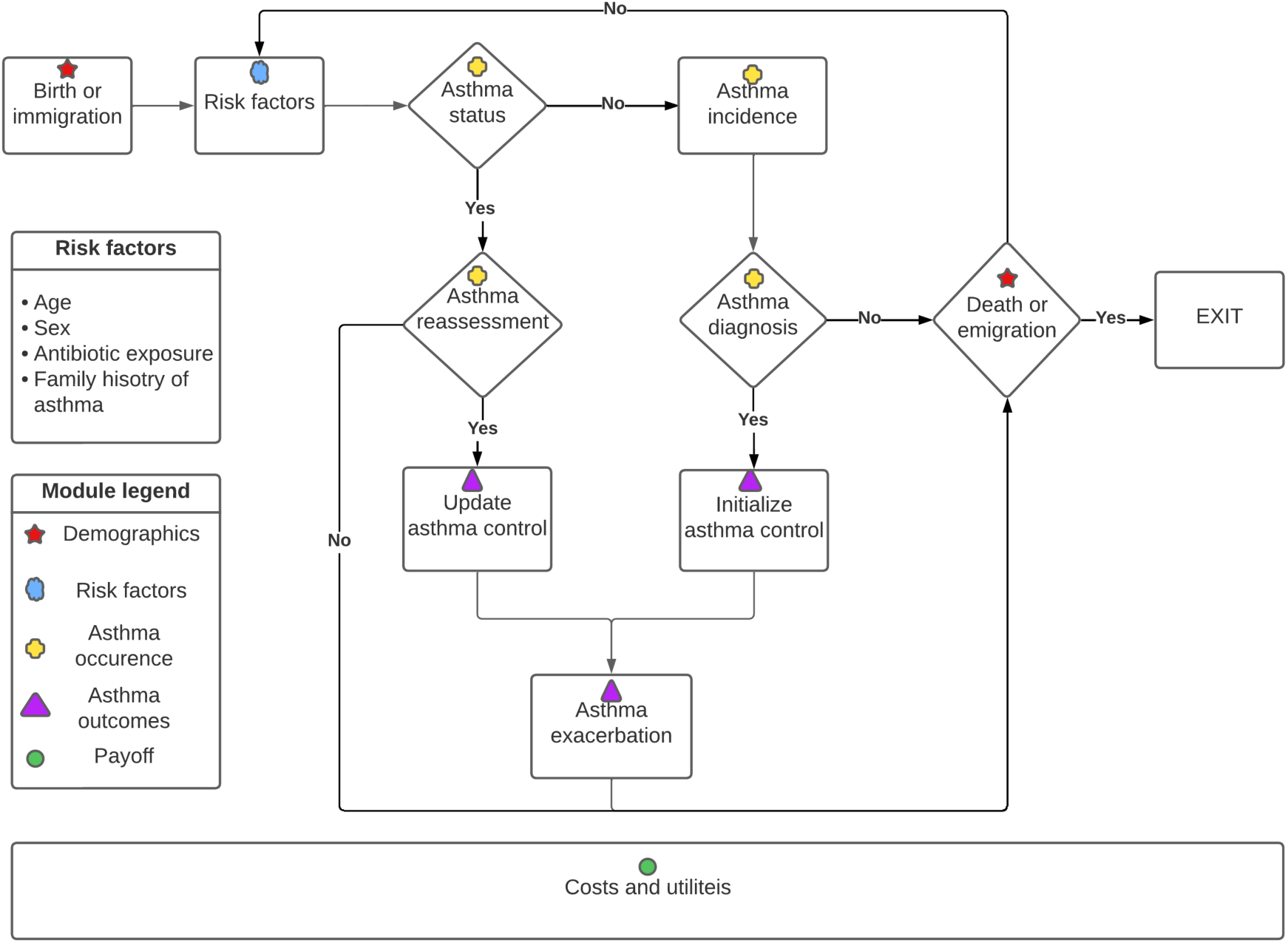
Schematic illustration of the reference asthma policy model.

**Table 1:** Structural equations by modules in the asthma model. The notation *x* ∗ *y* means main effects of *x* and *y* as well as their interaction effects. The notation 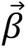 means a vector of regression coefficients of the necessary dimension. The notation poly(*x*, *q*) means polynomials of *x* up to degree *q* (i.e., *x*, *x*^2^, …, *x*^q^).

| Modules | Description | Source/Reference |
| --- | --- | --- |
| **Demographics** |  |  |
| Initial population | For a given year $\in \{2000, 2001, \dots, 2019\}$, the Statistics Canada estimated population size is used (treated as fixed). The empirical distribution of sex and age for the given year is used to assign age and sex of each individual $i$ in the initial population:<br>$(\text{sex}_i, \text{age}_i) \sim \text{Categorical}(p_{(\text{sex}, \text{age})} \vert \text{year})$ | Statistics Canada (2022a) |
| Birth | For a given year and projection scenario, the | Statistics Canada |
| | Statistics Canada estimated or projected number of births is used (treated as fixed). The empirical or projected distribution of sex at birth for the given year is used to assign sex of each birth $i$: $\text{sex}_i \sim \text{Bernoulli}(p_{\text{sex}}\vert \text{year}, \text{projection scenario})$ | (2022a,b) |
| Net immigration | For a given year and projection scenario, the number of net immigrants by sex and age is estimated by calibration (treated as fixed). The empirical or projected distribution by sex and age is used to assign sex and age of each net immigrant $i$: $(\text{sex}_i, \text{age}_i) \sim \text{Categorical}(p_{(\text{sex}, \text{age})}\vert \text{year})$ | Calibration |
| Net emigration | For a given year and projection scenario, the number of net emigrants by sex and age is estimated by calibration (treated as fixed). The empirical or projected proportion of net emigrants by sex and age is used to assign whether individual $i$ emigrates: $\text{emigration}_i \sim \text{Bernoulli}(p\vert \text{sex}, \text{age}, \text{year}, \text{projection scenario})$ | Calibration |
| Mortality | For a given year, sex, and age, the Statistics Canada estimated or projected life table is used to assign whether individual $i$ dies. $\text{death}_i \sim \text{Bernoulli}(p\vert \text{year}, \text{sex}, \text{age})$ | Canadian life table (Statistics Canada, 2022c) |
| **Risk factors** |  |  |
| Family history of asthma at birth ($X_1$) | Regardless of birth year, sex, and other characteristics, $X_1$ (the indicator of having parental history of asthma at birth) is generated: $X_1 \sim \text{Bernoulli}(p)$ | CHILD study (Takaro et al. 2015) |
| Infant antibiotic exposure ($X_2$) | For a given birth year and sex, $X_2$ (the number of courses of antibiotics individual $i$ receives in the first year of life) is generated by a Negative Binomial model. To prevent unrealistic extrapolation, the mean parameter is truncated at 0.05 (equivalent to 50 per 1,000 infants).<br>$X_2(\text{sex}_i, \text{birth year}_i) \sim \text{NegativeBinomial}(\max(0.05, \mu_i), \theta)$;<br>$\log(\mu_i\vert \text{sex}_i, \text{birth year}_i) = \beta_0 + \beta_1 \text{sex}_i + \vec{\beta}_2 \mathbf{1}[\text{birth year}_i > 2005] * \text{birth year}.$ | British Columbia Ministry of Health (2023) |
| **Asthma occurrence** |  |  |
| Effect of $X_1$ | The effect of $X_1$ in terms of log odds ratio (OR), denoted as $f_j(X_1, \text{age})$, on $j \in$ | Patrick et al. (2020) |
| | {asthma incidence (inc), asthma prevalence (prev)} is given by the following equation: $f_j(X_1, age)$<br>$:= \log(OR_j\vert X_1, age) = \mathbf{1}[age \geq 3 \text{ AND } X_1 = 1](\beta_{0j} + \beta_{1j}(\min(age, 5)) - 3).$ | |
| Effect of $X_2$ | The effect of $X_2$ in terms of log odds ratio (OR), denoted as $g_j(X_2, age)$, on $j \in \{\text{inc}, \text{prev}\}$ is given the following equation: $g_j(X_2, age)$<br>$:= \log(OR_j\vert X_2, age) = \mathbf{1}[(age \geq 3 \text{ AND } age \leq 7) \text{ AND } X_2 > 0](\beta_{0j} + \beta_{1j}age + \beta_{2j}\min(X_2, 3))$ | Lee et al. (2024) |
| Asthma prevalence $Y_{prev}$ | For a given year and individual characteristics, $Y_{prev}$ (the indicator of having an asthma label) is generated: $Y_{prev} \sim \text{Bernoulli}(p\vert \text{year}, \text{sex}, \text{age}, X_1, X_2)$<br>$\text{logit}(p\vert \text{year}, \text{sex}, \text{age}, X_1, X_2) = \beta_0 + \vec{\beta}_1 \text{poly}(\text{year}, 2) * \text{sex} * \text{poly}(\text{age}, 5) + f_{prev}(X_1, age) + g_{prev}(X_2, age)$<br>Note that the length of $\vec{\beta}_1$ is 34. For each of males and females, there are 2 main effects of year and year<sup>2</sup>, 5 main effects of age, age<sup>2</sup>, ..., age<sup>5</sup> and the corresponding 10 (first order) interaction effects between year and age. | British Columbia Ministry of Health (2023), Government of Canada (2023), calibration |
| Asthma incidence $Y_{inc}$ | For a given year and individual characteristics, $Y_{inc}$ (the indicator of developing an asthma label) is generated: $Y_{inc} \sim \text{Bernoulli}(p\vert \text{year}, \text{sex}, \text{age}, X_1, X_2)$<br>$\text{logit}(p\vert \text{year}, \text{sex}, \text{age}, X_1, X_2) = \beta_0 + \beta_1 \text{year} + \vec{\beta}_2 \text{sex} * \text{poly}(\text{age}, 5) + f_{inc}(X_1, age) + g_{inc}(X_2, age)$<br>Note that the length of $\vec{\beta}_2$ is 10. For each of males and females, there are 5 main effects of year, year<sup>2</sup>, ..., year<sup>5</sup>. | British Columbia Ministry of Health (2023), Government of Canada (2023), calibration |
| Asthma reassessment | For a given year, sex, and age, an individual labeled as asthmatic is reassessed. They maintain their asthma label with probability $p$, where $p$ is determined by calibration: asthma reassessment $\sim \text{Bernoulli}(p\vert \text{year}, \text{sex}, \text{age})$. | Calibration |
| **Asthma outcomes** |  |  |
| Asthma control | There are three levels of asthma control: 1 = uncontrolled (UC), 2 = partially controlled (PC), and | Economic Burden of Asthma (Chen et |
| | 3 = well-controlled (WC). For individual $i$ labeled as asthmatic, the proportion of time spent in each of the three asthma control levels in a year (conditional on their age and sex) is given by the probabilities generated by a random-effects ordinal regression model. For $Y_i$ the label of the asthma control level: $\text{logit}\left(P(Y_i \leq j \vert \text{sex}_i, \text{age}_i)\right) = \theta_j - (\beta_{0i} + \vec{\beta}_1 \text{sex}_i * \text{poly}(\text{age}, 2))$, for $j = 1, 2$, where $\beta_{0i} \sim N(0, \sigma^2)$ | al., 2013) |
| Asthma exacerbation | For a given year, sex, age, and the proportion of time spent in each of the control levels in that year (denoted $CT_i$ for $j = 1, 2, 3$), the number of exacerbations for an individual labeled as asthmatic is generated by a Poisson model:<br>number of exacerbations $\sim \text{Poisson}(\mu \vert CT_1, CT_2, CT_3, \text{year}, \text{sex}, \text{age})$.<br>$\log(\mu \vert CT_1, CT_2, CT_3, \text{year}, \text{sex}, \text{age}) = \beta_0 + \sum_j \beta_j CT_j$, where $\beta_0$ is determined by calibration as a function of year, sex, and age. | Economic Burden of Asthma (Chen et al., 2013), the GOAL study (Bateman et al., 2004), Lee et al. (2022a), calibration |
| Exacerbation severity | There are four levels of exacerbation severity: 1 = mild, 2 = moderate, 3 = severe, and 4 = very severe. Given the number of exacerbations $N_i$ in a year and the past history of very severe exacerbations $n_{i4}^{\text{past}}$, the number of exacerbations $n_{ij}$ in each severity level $j$ is simulated for each individual $i$ by the Dirichlet-Multinomial distribution. A preliminary vector of probabilities for the severity levels is sampled from a Dirichlet distribution: $\mathbf{w}_i^{\text{pre}} \sim \text{Dirichlet}(\boldsymbol{\alpha})$. If individual $i$ experienced any previous very severe exacerbations, their probability of very severe exacerbations is increased to yield: $\mathbf{w}_i = \mathbf{w}_i^{\text{pre}}(n_{i4}^{\text{past}})$ (see the main text for details). The number of exacerbations by the severity levels is generated by a multinomial distribution: $(n_{i1}, n_{i2}, n_{i3}, n_{i4}) \sim \text{Multinomial}(N_i, \mathbf{w}_i)$. | Yaghoubi et al. (2020), Bateman et al. (2018), Lee et al. (2022b) |
| **Payoffs** |  |  |
| Baseline utility | The utility values measured in quality-adjusted life years, ranging from 0 (death) to 1 (perfectly healthy) based on EQ-5D are used to assign the base utility of each individual by sex and age. | Yan et al. (2024) |
| Disutility due to asthma control (per year) | The base utility value of an individual labeled as asthmatic decreases based on their asthma control status. Let $du_j^C$ denote the disutility due to having asthma control level $j$ in a year. Then the total disutility due to asthma control is given by: $\sum_j du_j^C CT_j$ | Einarson et al. (2015) |
| Disutility due to asthma exacerbation (per year) | The base utility value of an individual labeled as asthmatic decreases if they experience asthma exacerbations. Let $du_j^E$ denote the disutility due to experiencing a single asthma exacerbation of severity $j$ in a year and $n_j$ be the number of exacerbations of severity $j$ for $j = \text{mild, moderate, severe, and very severe}$. Then the total disutility due to asthma exacerbations is given by: $\sum_j du_j^E n_j$. | Aldington and Beasley (2007), Yaghoubi et al. (2020), Einarson et al. (2015) |
| Direct annual costs due to asthma control | Let $c_j^C$ be the direct annual costs due to asthma control level $j$. Then the total cost due to asthma control is given by: $\sum_j c_j^C CT_j$. | Yaghoubi et al. (2020) |
| Direct annual costs due to asthma exacerbation | Let $c_j^E$ be the direct annual costs due to one asthma exacerbation of severity $j$. Then the total cost due to asthma exacerbations in a year is given by: $\sum_j c_j^E n_j$ | Yaghoubi et al. (2020) |

We describe two high-level assumptions in the model. We assumed that asthma was not labeled for children under 3 years of age due to difficulty with performing and confirming assessment for this age group. As such, children under this age were not assigned any asthma attributes. Further, we assumed that death or emigration occurred at the end of the time cycles. Put differently, any events they experience in the year they die (or emigrate) were assumed to happen before their death (or emigration).

In the remainder of this section, we describe each of the five modules in detail and then provide an overview of how these modules are connected including pseudocode of the microsimulation model at the end.

### Demographics module

The demographics module consists of birth, immigration, emigration and mortality equations. At the start of the simulation, an initial population is generated for the specified base year. In subsequent years, virtual individuals enter the simulated population through birth or immigration according to the estimates or projections of population growth and aging, and exit the simulated population when one of the following events occur: death, emigration, or reaching the end of the time horizon.

To model the initial population prior and up to 2019, we used the population estimates by sex and age from Statistics Canada (Statistics Canada, 2022a). To model the initial population for later years, we applied a population projection by sex and age from Statistics Canada (Statistics Canada, 2022b). Among the nine population projection scenarios provided by Statistics Canada (low growth, five levels of medium growth, high growth, slow aging, and fast aging) (Statistics Canada, 2022b), we found that the third medium (M3) growth resulted in the lowest root mean squared error of the total population at the national level when compared with the observed data in 2020 and 2021 (Appendix Section 1). As such, the demographics module was calibrated to the M3 growth.

Birth is one of the two ways for individuals to enter the simulation after the initialization of the population. The number of births by sex and year was based on the estimate (2000-2019) or projection (2020-2065) from Statistics Canada (Statistics Canada, 2022a,b). Statistics Canada does not provide a breakdown of immigrants and emigrants by sex and age in their population estimates and projections. We decided to model the *net* immigrants and *net* emigrants via model calibration. For each year, we calculated the number of individuals required to immigrate and emigrate, by sex and age, to match the estimated or projected size of the population. Specifically for each year, we computed the total numbers of net immigrants and net emigrants by summing over all combinations of sex and age and treated these totals as non-random. We then computed the empirical distribution of net immigrants by sex and age by dividing the count of each sex–age cell by the total number of net immigrants for each year. For net emigrants, we calculated the probability of emigrating out of the country by sex and age by dividing the count of each sex–age cell by the total number in the population for each year. We then used these empirical distributions to assign sex and age of each immigrant (via a categorical distribution) and to determine whether an individual emigrates (via a Bernoulli distribution), respectively.

In the current version, the mortality rate was not differentiated between individuals with asthma and those without asthma, as deaths due to asthma are very rare (O’Byrne et al., 2019). In Canada, asthma was responsible for a very small proportion of all-cause deaths annually between 2000 and 2020 (the maximum value was less than 0.14%) (Statistics Canada, 2022d). As such, differentiation was not deemed necessary.

For mortality, we used the estimated life tables to model whether an individual dies at the end of each time cycle (Statistics Canada, 2022c). However, Statistics Canada does not provide a projected life table but a predicted life expectancy of an individual at birth in year 2068 for each projection scenario. For M3, this value is 87.0 years for males and 90.1 years for females (Statistics Canada, 2022b). To reflect this increase in life expectancy, we modified the latest life table (2020) for each sex by calibrating the probability of death across all ages as follows:

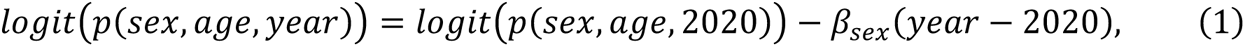

where *p*(*sex*, *age*, 2020) is the probability of death from the 2020 life table. Our goal is to find β_*sex*_ such that the projected life table matches the 2068 life expectancy predicted by Statistics Canada. Given β_*sex*_, we can calculate the probability of death for each age in 2068 using (1) and compute the corresponding projected life expectancy at birth (details are provided in Appendix Section 2). We solved for β_*sex*_ by minimizing the absolute error between the projected and targeted life expectancy values using a bisection method (uniroot function in R).

### Risk factor module

We describe here how each risk factor is instantiated for a virtual individual. In the current version of the model, the following risk factors are included: age, biological sex, family history of asthma at birth, and infant (< 1 year of age) antibiotic use. Age and sex of a virtual individual were determined by the demographics module. The probability of having family history of asthma at birth was assumed to be constant across birth year, age, and sex. To obtain an estimate of the probability of having family history of asthma at birth, we used the Canadian Healthy Infant Longitudinal Development (CHILD) study, an ongoing representative birth cohort of 3,455 families (Takaro et al., 2015). We found that 29.3% (95%CI: 27.3–31.1) of the children in the study had parental asthma. We assumed that all virtual individuals had this probability of having family history of asthma at birth, regardless of birth year, age, and sex.

To obtain an estimate of the infant antibiotic exposure, we used the antibiotic prescription rate as a proxy. Using the prescription data from the British Columbia Ministry of Health for the period from 2000 to 2019 (British Columbia Ministry of Health [creator], 2023), we fitted a negative binomial model with the log link and the number of courses of antibiotics in the first year of life as the response variable, birth year and biological sex as linear covariates, and the logarithm of the number of new births as an offset. In addition, we added an interaction term between birth year and an indicator for the introduction of an antibiotic stewardship program in British Columbia in 2005 (Mamun et al., 2019).

We observed a decreasing trend of antibiotic exposure, with males receiving more courses of antibiotics than females. After the antibiotic stewardship program was introduced in 2005, the trend became steeper (Figure 3). For subsequent years we assumed the stewardship program would remain in place and the corresponding trends were extrapolated. However, extrapolation eventually leads to near zero rates of antibiotic exposure which is unrealistic. To prevent this, we assumed a minimum rate of 50 (per 1,000 persons) after consultation with the steering committee. We used this truncated rate parameter in the negative binomial model to simulate the number of antibiotic prescriptions in the first year of life for each virtual individual. For individuals born prior to 2001, we treated their birth years as 2001 for simulating their number of antibiotic prescriptions (i.e., we did not use the model to extrapolate into the past).

**Figure 3:**
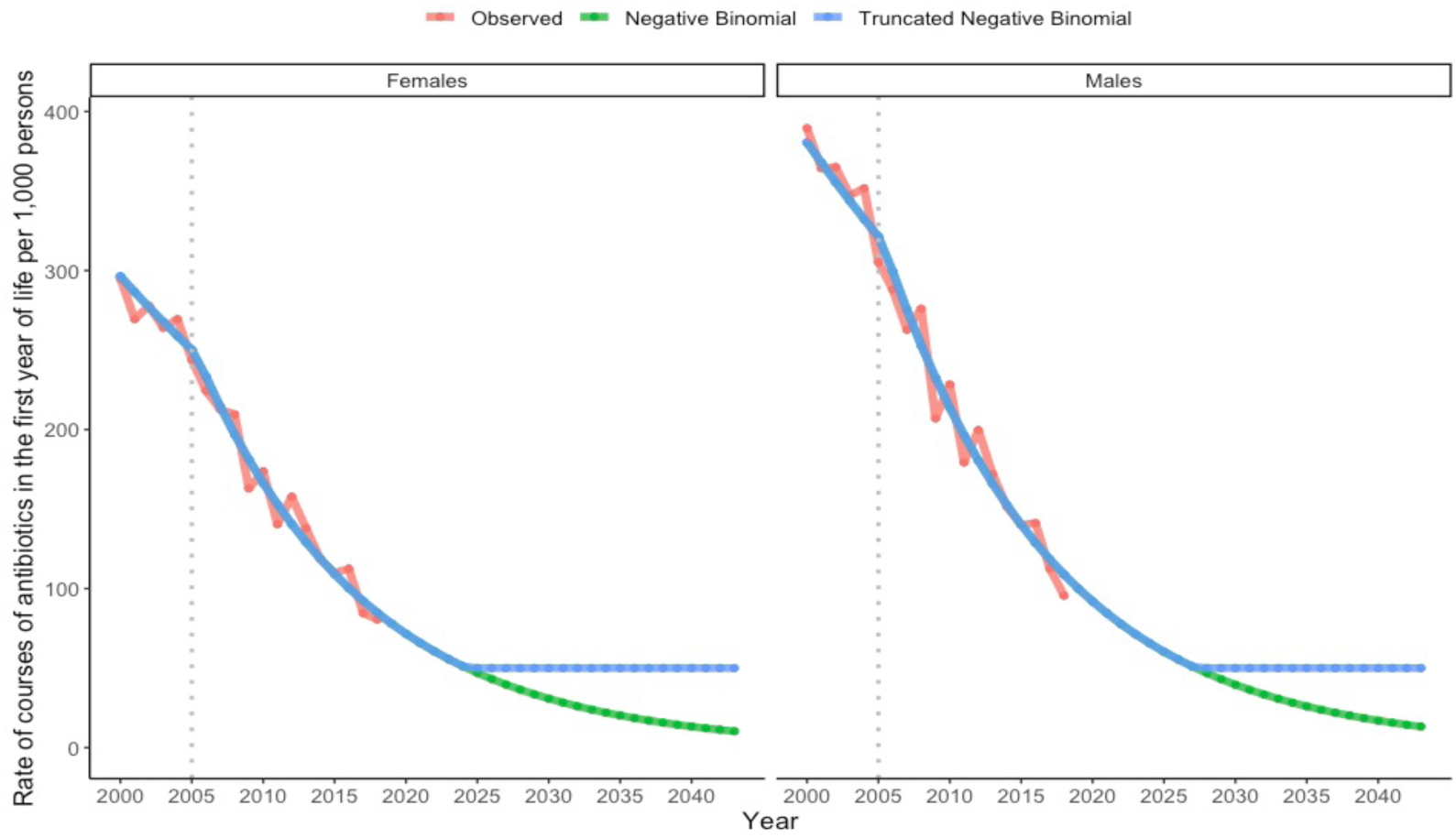
Trends in the rate of courses of antibiotics in the first year of life in British Columbia by sex (dotted grey: 2005; red: observed; green: predicted; blue: truncated).

### Asthma occurrence module

This module is responsible for asthma labeling. Recall that we decided to model labeled asthma states rather than true asthma states. For the initial population, we need a prevalence equation to determine whether an incoming virtual individual is labeled as asthmatic in the base year of the microsimulation. For individuals not labeled as asthmatic for each time cycle in the model, we need an incidence equation to determine whether they become labeled as asthmatic. We assumed that immigrants have the same asthma incidence and prevalence rates as Canadians. Recall that asthma was not modelled for children under 3 years of age, as it is difficult to perform tests to perform and confirm asthma assessment for that age group (Jones et al., 2019). Accordingly, we did not assign any asthma attribute to children under 3 years of age. The equations for incidence and prevalence are provided in Table 1.

Values for the parameters in these equations were obtained in a stepwise fashion. We started with population-based age– and sex-specific ‘crude’ prevalence and incidence without the effect of other risk factors (family history of asthma at birth and exposure to antibiotics in the first year of life). Then we introduced asthma reassessment to calibrate the age– and sex-specific crude incidence and prevalence for each year. Finally, we incorporated and calibrated for the effect of the two remaining risk factors.

### Estimation of crude asthma prevalence and incidence

In this subsection, we explain how we obtained estimates of crude asthma prevalence and incidence at the national level. For this purpose, we used two data sources: a national survey (Government of Canada, 2023) and an administrative database of British Columbia (BC). The Canadian Community Health Survey (CCHS) is a cross-sectional self-reported survey for 12 years or older at the national level. It provides asthma prevalence estimates at the national level but does not report on asthma prevalence for the age group of less than 12 years of age or on asthma incidence for any age. On the other hand, both asthma prevalence and incidence rates in BC were provided in the administrative data. However, in the administrative data, asthma labeling was made with healthcare utilization based on the diagnostic codes (ICD codes–10-CA: J45; ICD-9-CA: 493): one or more asthma-related hospitalization, OR two or more physician visits within one year, OR one or more physician visits and two or more asthma prescriptions within one year (for a list of asthma prescriptions, see Ministry of Health (2022)).

Upon examining asthma prevalence in the CCHS data, we found that prevalence did not differ much between BC and Canada (green and blue lines, respectively in Figure 4). On the other hand, there was a considerable discrepancy in asthma prevalence between the CCHS and BC administrative data (green and red line, respectively in Figure 4). As CCHS was a self-reported survey which is subject to self-selection and response bias, we deemed the estimates from the BC administrative data to be more reliable. As such, we made a simplifying assumption that nationwide asthma prevalence and incidence rates were equal to those from the BC administrative data.

**Figure 4:**
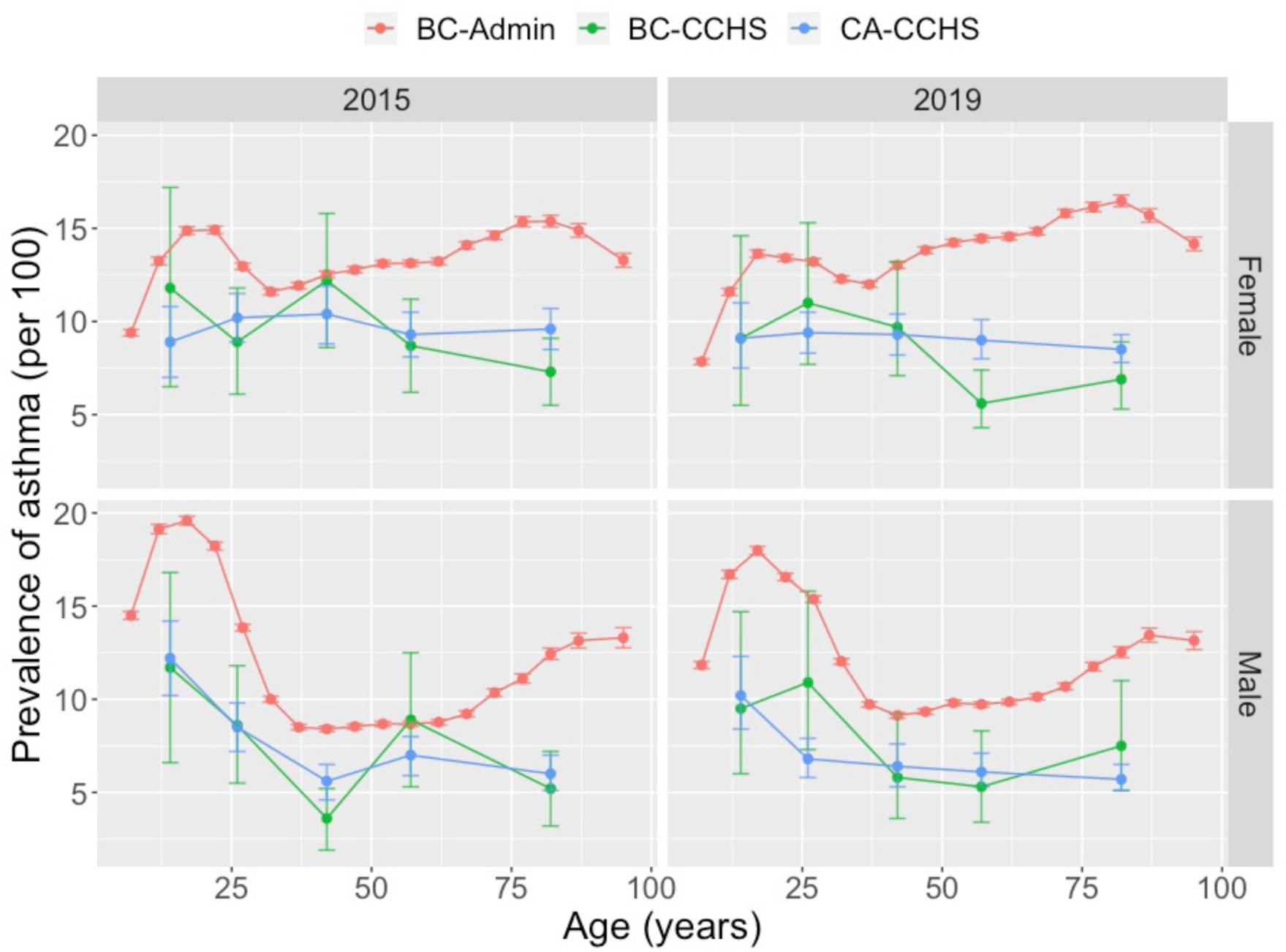
Observed asthma prevalence across age in 2015 and 2019 for British Columbia (BC) and Canada (CA) based on the BC administrative data (red) and CCHS data (green: BC; blue: CA) by sex. CCHS: Canadian Community Health Survey.

In the BC administrative data, we had information on observed prevalence and incidence from 2000 to 2019, stratified by 5-year age bands (0-4, 5-9, …, 84-89, 90+ years), biological sex, and calendar year(British Columbia Ministry of Health [creator], 2023). To estimate the prevalence and incidence for each age bin, we first took the mid-point of the five-year age bands as the age for the corresponding prevalence and incidence. We discarded data on age > 65 years due to potential mislabeling among older population and assumed that the incidence and prevalence rates for age > 63 years (the mid-point of the last age band) remained constant at the rates for 63 years of age. For asthma incidence, we fitted a linear regression model with the log of the incidence rate as a linear combination of time, sex, poly(age,5) (recall that poly(*x*, *q*) stands for the polynomials of *x* up to degree *q*), as well as interaction terms of sex and poly(age,5) (Figure 5). For asthma prevalence, we fitted a linear regression model with the log of the prevalence rate as a linear combination of poly(time,2), sex, poly(age,5) and all their interactions (Figure 6).

**Figure 5:**
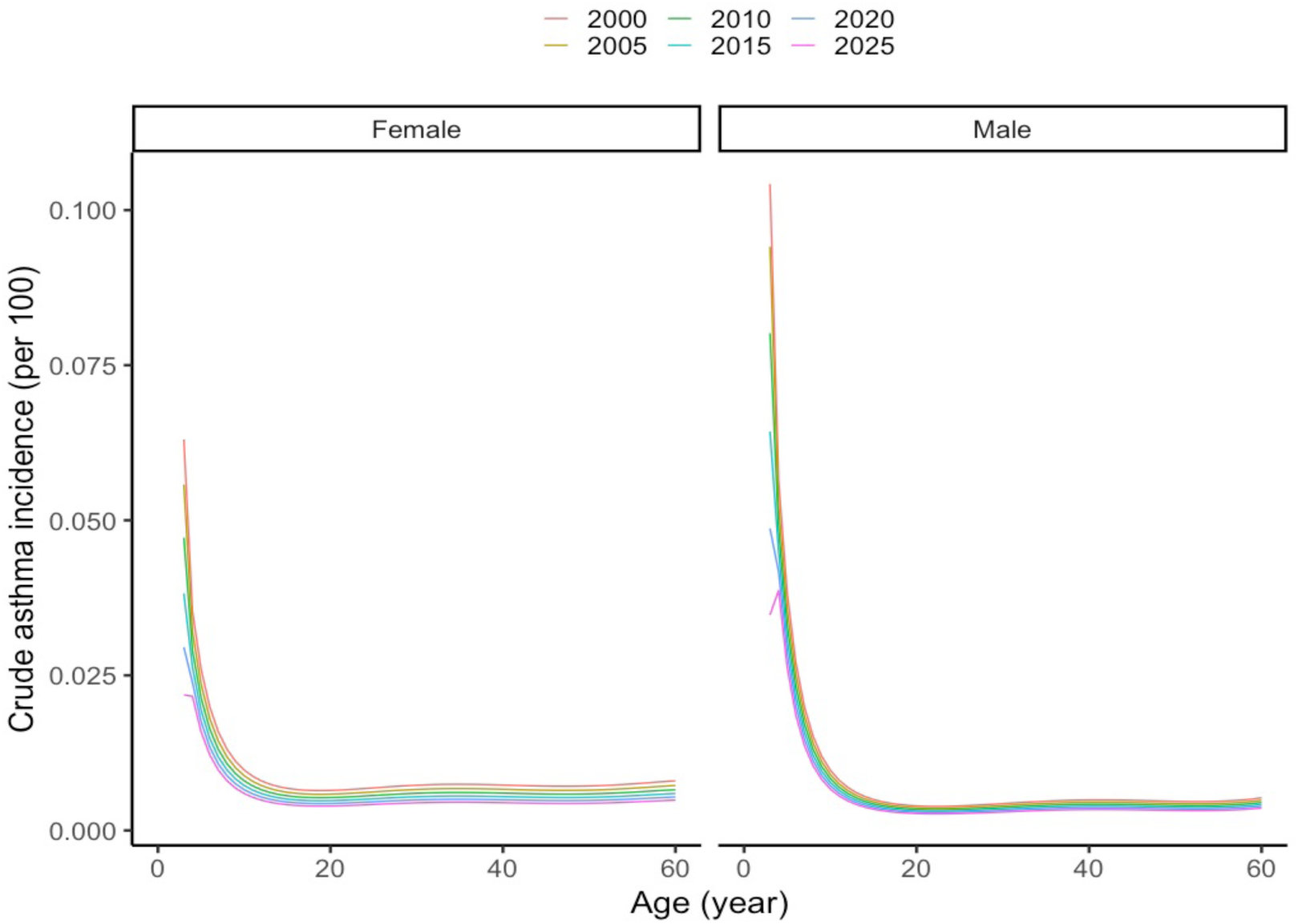
Estimated asthma incidence for selected years by sex using the administrative data of British Columbia.

**Figure 6:**
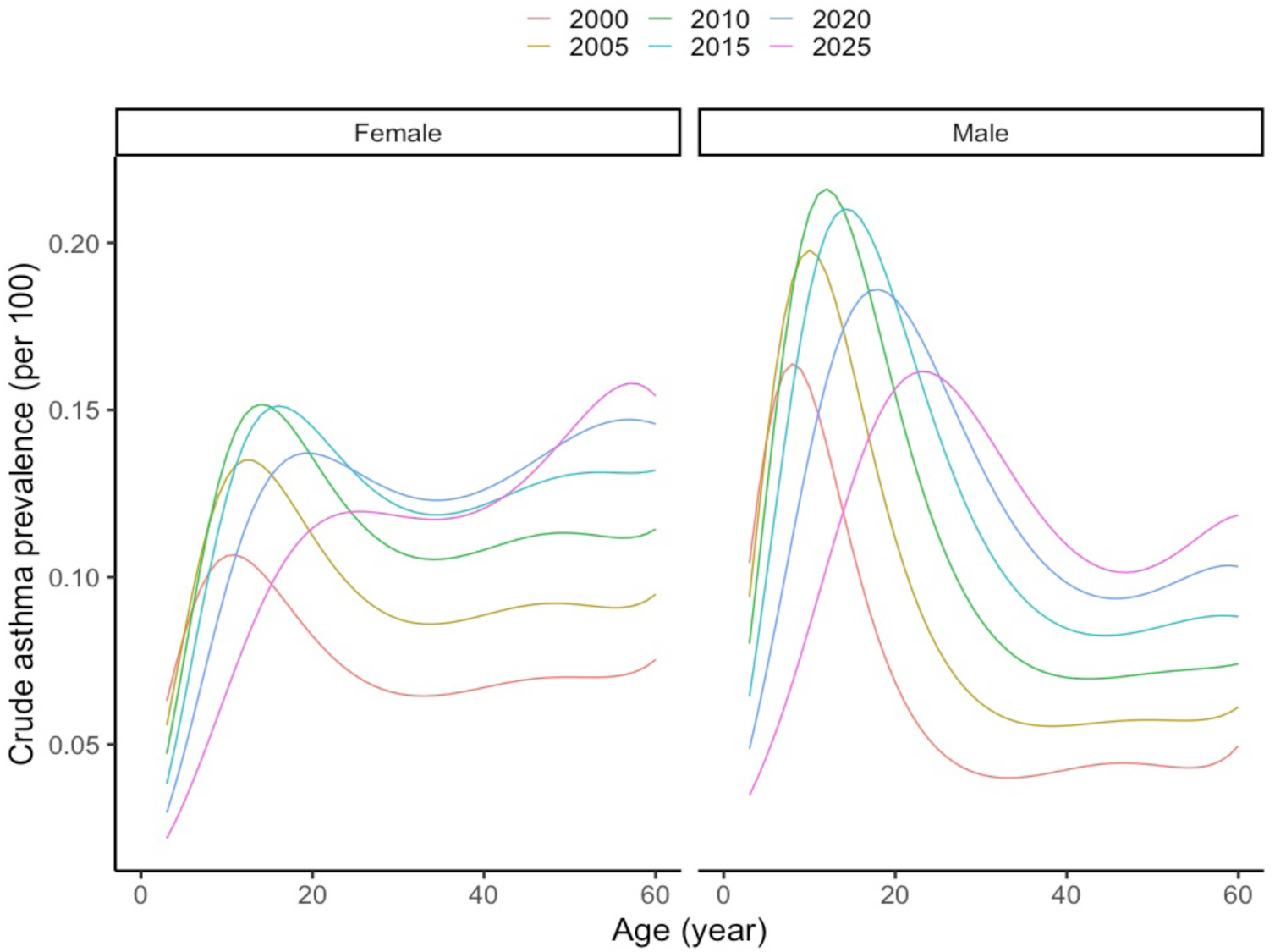
Estimated asthma prevalence for selected years by sex using the administrative data of British Columbia.

For the past (prior to 2000), we assumed the estimates of prevalence and incidence from 2000. For the future (2020 onwards), we assumed that current incidence and prevalence trends continued up to 2025, and stayed constant thereafter. For the initial population and immigrants, the asthma prevalence estimates would be used to assign the asthma label to all individuals 3 or more years of age. For individuals not labeled as asthmatic, the asthma incidence estimates (except for individuals less than 3 years of age) would be used to simulate whether an individual becomes labeled as asthmatic in each time cycle. Of note, this implies we assume that immigrants have the same asthma incidence and prevalence rates as Canadians.

While these crude asthma prevalence and incidence estimates could be used to simulate asthma cases in the asthma model, the asthma model would generate excess asthma cases without accounting for asthma dormancy or remission (e.g., the asthma prevalence rises from early childhood to young adulthood but then falls). In addition, we still need to incorporate risk factors. In the next section, we describe how we calibrated the crude asthma and incidence estimates through the asthma reassessment submodule before incorporating the risk factors.

### Reassessment

Asthma prevalence varies across age, characterized by a hump in early age. Asthma is not curable and from a biologic perspective, its core pathology does not disappear (Thomas et al., 2022). Such variation is instead explained by the ‘dormancy’ of asthma, a prolonged period with no asthma symptoms. Such dormancy might be endogenous (e.g., outgrowing of asthma from childhood to adulthood) or exogenous (e.g., change in environment and risk of asthma triggers) (Bisgaard and Bønnelykke, 2010). As well, in the community, misdiagnosis of asthma occurs mostly due to confounding by other allergic diseases or respiratory diseases (Kavanagh et al., 2019), which can also explain reduction in the prevalence of asthma by age. Correspondingly, we modeled the clinical status of asthma by a sub-module, asthma reassessment, to evaluate whether an individual labeled as asthmatic remains labeled as asthmatic.

We assume the following relationship among asthma incidence, prevalence, and reassessment. For specific sex and age, the number of individuals labeled as asthmatic at the current year *t* consists of individuals labeled as asthmatic from the past year (*t* − 1) who remain labeled as asthmatic and individuals without an asthma label from the past year who become labeled as asthmatic in the current year. For specific sex and age (arguments dropped for brevity), let *prev*_*t*_ be the probability (or equivalently the proportion) of being labeled as asthmatic in year *t*, *inc*_*t*_ be the probability of becoming labeled as asthmatic, and *p*_*t*_ (*reassessment*) be the probability of maintaining the asthma label. Of note, we do not have to consider immigrants, emigrants, or death as we assume that death or emigration occurs at the end of the year and that immigrants have the same prevalence and incidence as the Canadian population. Then the relationship can be described as follows:

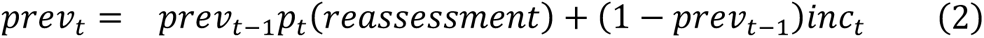

We solved for *p*_*t*_ (*reassessment*) for each year, sex, and age, as all the other quantities were known from the previous section. In the microsimulation, an individual is labeled as asthmatic based on the asthma prevalence equation when they enter the simulation. In a subsequent time cycle, if they are labeled as asthmatic, then their label is reassessed and maintained with *p*_*t*_ (*reassessment*). If not, their asthma label is determined by the asthma incidence equation.

### Incorporating the effect of risk factors

In this subsection, we describe how the effect of the risk factors, namely family history of asthma at birth and infant (< 1 year of age) antibiotic exposure, are estimated and incorporated into the asthma incidence and prevalence equations.

Using the CHILD study data, Patrick et al. (2020) found that family history of asthma was associated with an increase in the risk of being labeled as asthmatic. They used a multivariable logistic regression to establish the association with the prevalence of asthma at the age of 3 years with an odds-ratio [OR] of 1.13 (95%CI: 0.66–1.95) and at the age of 5 years with an OR of 2.40 (95%CI: 1.13–5.09). In addition to sex, they adjusted for presence of older siblings, exposure to antibiotics in the first year of life, ethnicity, mode of delivery, birth weight, parental asthma, breastfeeding, exposure to tobacco smoke, season of birth, and exposure to environmental nitrogen oxide (these additional risk factors should be incorporated in the future). We used linear interpolation on the log OR scale to estimate the risk at the age of 4 years, and for age greater than 5 years, we assumed that the risk stayed constant at the level for 5 years of age. That is, the log odds ratio for having family history of asthma at birth (for age ≥ 3 years) can be expressed as:

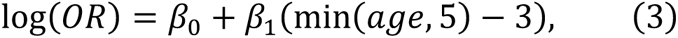

where β_0_ = log(1.13) and β_1’_ = 1log(2.40) − log(1.13)2/2. This equation corresponds to *f_i_*(⋅) (“Effect of *X*_’_”) in Table 1.

A recent systematic review found support for the association between exposure to antibiotics in early life and the risk of being labeled as asthmatic (Duong et al., 2022). However, the association is mostly attributable to exposure to antibiotics in the first year of life (Hoskinson et al., 2023). In consultation with the steering committee, we assumed an association only for the first year of life. To obtain an estimate of the age-specific dose-response of the number of courses of antibiotics in the first year of life on the risk of being labeled as asthmatic, Lee et al. (2024) carried out a meta-analysis using the summarized data from the systematic review. Applying strict inclusion-exclusion criteria, they included six studies that reported on the dose response relationship, including a Canadian study by Patrick et al. (2020), with low risk of bias. They fitted a random-effects meta-regression model with the number of courses of antibiotics (0, 1, 2, 3, 4, 5 or more) and age of asthma diagnosis as covariates. They found a detrimental dosage effect up to 7 years of age and a diminishing effect with age of antibiotic exposure in the first year of life. We followed their assumption of no effect of the exposure to antibiotics in the first year on the risk of asthma beyond 7 years of age.

We made a further assumption to facilitate the calibration (discussed next). As the antibiotic prescription rate was not high, the probability of having more than three courses of antibiotics in the first year of life was practically zero. Hence, we reclassified the levels 3, 4, and 5+ together as one level, 3+, and the log OR for 3+ was assumed to be equal to the log OR for 3. That is, the log odds ratio for having any number of courses of antibiotics (dose) in the first year of life (for age ∈ {3, …, 7} years and dose > 0) can be expressed as:

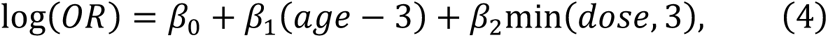

where β_0_ = 1.826, β_’1_ = −0.225, and β_2_ = 0.053. This equation corresponds to *g_j_*(⋅) (“Effect of *X*_1_”) in Table 1.

Now incorporating the effect of risk factors into the asthma incidence and prevalence equations requires calibration, so that the marginal asthma prevalence (i.e., the sum of the products of the prevalence of each combination of the risk factors and the corresponding asthma prevalence) remains calibrated.

At calendar year *t*, sex *s*, and age *a*, let *X*(*t*, *s*, *a*) = 0,1,2, …, *q* be a categorical risk factor that takes levels *x* = 0,1, …, *q*. For brevity, we drop the arguments *t*, *s*, *a* in the following text. Let *p_target_* be the target marginal asthma prevalence, *OR_x_* be the association between the risk factor level *x* and asthma labeling, and *p_x_* be the prevalence of the risk factor level. With *p_prev_* denoting the asthma prevalence at level *x* and 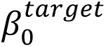 denoting the logit of the target asthma prevalence, we seek a correction term δ for the intercept in the following asthma prevalence equation

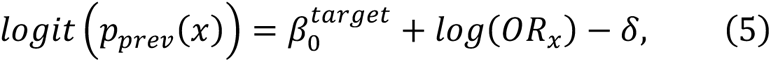

such that the marginal asthma prevalence is calibrated (i.e., 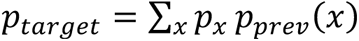) while maintaining the ORs. There is a unique solution to this optimization problem (monotone decreasing in δ), and we used the Broyden–Fletcher–Goldfarb–Shanno (BFGS) algorithm to solve for δ (Fletcher, 2000). Multiple categorical risk factors can be represented as a single categorical risk factor, and the same method can be applied.

To obtain the calibrated asthma prevalence equation in Table 1, we took the following steps for each year, sex, and age. First, we represented the logit of the crude asthma prevalence by 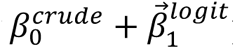 poly(year,2) ∗ sex ∗ poly(age,5). These terms represent 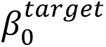 in Equation 5. The effect of level *x* of the risk factors *X*_1’_ and *X*_2_is incorporated with using *f_prev_* and *g_prev_*, which represents log(*OR_x_*) in Equation 5. Finally, we evaluated 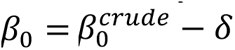 by β_0_ to obtain the final form of the calibrated equation.

Calibration for the asthma incidence equation is more complicated since the effects of the risk factors are unknown and need to be estimated such that both the marginal asthma incidence and the ORs in the asthma prevalence equation remain unchanged. We assumed that the same form of relationship between the risk factors and asthma prevalence for asthma incidence (*f_j_* and *g_j_* in Table 1) and that the risk factor equations for asthma prevalence and incidence are equal at age of 3 years (i.e., *f_inc_* = *f_prev_* and *g_inc_* = *g_prev_* at age of 3 years). In other words, the values of β_0_ in *f_inc_*, and β_0_ and β_2_in *g_inc_* are equal to those of *f_prev_* and *g_prev_*, respectively. Our goal is then to optimize β_1’_’s in *f_inc_* and *g*_inc_.

Suppose we start with an initial guess of β_1’_’s in *f*_i*nc*_ and *g_inc_* (we used the corresponding values in *f_prev_* and *g_prev_*). We apply the calibration method used for the asthma prevalence equation to adjust for the marginal asthma incidence, giving us the adjusted asthma incidence for each combination of levels of the risk factors. For each combination of levels (excluding the reference level), the first step is to construct a 2×2 contingency table for labeled asthmatic by level *x* versus the reference level from the previous time cycle. The second step is to construct the contingency table for the current time cycle using the asthma incidence equation and asthma reassessment. The third step is to compute the resulting ORs for asthma prevalence and compare them with the ORs from the asthma prevalence equation. The absolute differences are aggregated over all combinations of year (up to the stabilization year, 2025), sex, and age. This process is iterated until convergence with the BFGS optimization algorithm. Details are provided in Appendix Section 3.

In summary, we have described how we estimated each parameter in the asthma incidence and prevalence equations. In the next section, we explain the asthma outcomes module that an individual labeled as asthmatic experiences in the microsimulation.

### Asthma outcomes module

Two main features of the disease course of asthma are asthma control and asthma exacerbation. Asthma control refers to how well asthma and the risk of adverse outcomes can be managed with risk factor modifications or treatment (Global Initiative for Asthma, 2023). There are different methods for the assessment of asthma control, with the definition adopted by the Global Initiative for Asthma (GINA) being the most popular.

According to this definition (Global Initiative for Asthma, 2023), at any given time, an individual with asthma is either uncontrolled (UC), partially controlled (PC), or well-controlled (WC). Asthma control is evaluated based on the frequency of asthma symptoms (e.g., night waking due to asthma).

An asthma exacerbation (or a ‘flare-up’) is a sudden worsening of asthma symptoms, such as wheezing, coughing, and chest tightness (Global Initiative for Asthma, 2023).

Contemporary management of asthma emphasizes reducing the risk of asthma exacerbations. Moreover, asthma exacerbations are a key variable for deciding a treatment strategy. As asthma control is determined by assessing symptoms, exacerbation rate is naturally associated with asthma control.

### Asthma control

To estimate the control levels in Canada, we used the data from the Economic Burden of Asthma (EBA) study (Chen et al., 2013). EBA was a prospective representative observational study of 618 participants aged 1-85 years (74% are 18 years or older) with self-reported, physician diagnosed asthma from BC. Based on the GINA guidelines, asthma control and numbers of exacerbations were measured every 3 months for a year. For the asthma control data, our goal was to fit a model for generating the proportion of time that an individual labeled as asthmatic spends in each control level. We fitted a random-effects ordinal regression model with the logit link and included age, age squared, sex, and their interactions as fixed-effects and individuals as a random-effect using the clmm function from the *ordinal* R package (Christensen, 2023). While predictions from this model are the probabilities of being in each of the control levels during the 3-month period, we assumed them to be the proportion of time spent in the control levels. Further, we assumed that those predictions apply for one year instead, matching the time cycle unit of the simulation, and that there was no time trend as well as no dependency on the past history of asthma control and exacerbations. In short, for each virtual individual labeled as asthmatic, we sampled an individual-specific intercept from the estimated distribution of the random-effects, and with that intercept in the asthma control prediction model, we simulated the proportion of time spent in each of the control levels in each time cycle.

### Asthma exacerbation

For each individual labeled as asthmatic in each year, we used a Poisson regression model to simulate the number of exacerbations in each cycle conditional on the proportion of time spent in each of the asthma control levels by combining evidence from the EBA study (Chen et al., 2013) and the Gaining Optimal Asthma controL (GOAL) study (Bateman et al., 2004). Combining these two sources was deemed necessary, because the EBA study alone did not have enough events for robust estimation of exacerbation rate. The GOAL study was a one-year randomized, double-blind clinical trial with 3,421 individuals aged between 12 and 80 years with uncontrolled asthma at entry to the study, with asthma exacerbations as the primary outcome. We acknowledge a potential limitation of this study as the results may not be generalizable to the general asthmatic population. The cohort is not likely representative of the general population (individuals were uncontrolled at entry) and the treatment escalation strategy is likely to differ in practice.

We first obtained the annual rate of exacerbation from the EBA study, as 0.347/year, as well as the proportion of time spent in the control levels, well-controlled (WC) = 0.340, partially-controlled (PC) = 0.474, and uncontrolled (UC) = 0.186. An analysis of the GOAL study provided the (rounded) annual exacerbation rates for each asthma control level: rate(exacerbation|WC) = 0.1, rate(exacerbation|PC) = 0.2, and rate(exacerbation|UC) = 0.3. Thus, we assumed that the annual exacerbation rate for an PC/UC asthmatic individual is twice/thrice higher than for a WC asthmatic individual. Using the annual exacerbation rate from the EBA study, we solved for rate(exacerbation|WC) in the relationship rate(exacerbation) = P(WC) * rate(exacerbation|WC) + P(PC) * rate(exacerbation|PC) + P(UC) * rate(exacerbation|UC) to obtain the annual exacerbation rate conditional on each asthma control level: rate(exacerbation|WC) = 0.188, rate(exacerbation|PC) = 0.376, rate(exacerbation|UC) = 0.564.

We simulated the total number of exacerbations in a year experienced by an asthmatic individual as Poisson, where the log of the mean parameter is a linear function of the time spent in each of the asthma control levels *CT_j_*, with the corresponding coefficient estimates, ∑*_j_* β*_j_ CT_j_* (e.g., log(0.188) for *CT*_3_), and a calibration term β_0_. Estimation of the calibration term is described in the subsection, “Calibration for the exacerbation module’’ below after the subsections on exacerbation severity and initialization of the exacerbation module.

### Exacerbation severity

Asthma exacerbation is commonly classified as mild, moderate, or severe (Global Initiative for Asthma, 2023). We added a fourth level, *very severe*, that requires hospitalization (Castillo et al., 2017). To assign the severity of exacerbation, we used data from the Symbicort Given as Needed in Mild Asthma (SYGMA) II study, a double-blind multi-center clinical trial with individuals with mild asthma (n=4,176) (Bateman et al., 2018). The proportion of exacerbation by severity was 49.5% for mild, 19.5% for moderate, 28.3% for severe and 2.6% for very severe (Bateman et al., 2018; Yaghoubi et al., 2020). We used those values as the shape parameter α for a Dirichlet distribution to generate a preliminary vector of probabilities **W***^pre^*, for the severity levels. We then incorporated the effect of very severe exacerbations on subsequent very severe exacerbations (Lee et al., 2022b). If an individual labeled as asthmatic had a past history of very severe exacerbation, the probability for the very severe level increased by a factor of 1.79 if less than 14 years of age and 2.88 otherwise. The probabilities for the other levels were correspondingly scaled down such that the sum of the probabilities was 1. Finally, given the total number of exacerbations for an individual labeled as asthmatic in a year and the final probability vector **W** for the severity levels, we generated the number of exacerbations in each severity level with a multinomial distribution.

### Initialization of the exacerbation module

We needed to assign whether any very severe exacerbations were previously experienced by each individual labeled as asthmatic in a prevalent population. To do so, we first needed to determine the number of time cycles that individual was labeled as asthmatic since asthma can be reversible. Calculating this was computationally burdensome due to an explosion of possible states. To simplify, we made the assumption that asthma was not reversible in these individuals and ran a mini-simulation to determine the time cycle the individual was labeled as asthmatic and then calculated the probability of having no very severe exacerbations from that incidence to the present. Finally, we tossed a coin to determine whether this individual had at least one very severe exacerbation. Our simplifying assumption implies the number of years that the individual was labeled as asthmatic was overestimated. However, given the low chance of asthma reversibility in our dataset, we posited that overestimation was not too severe.

### Calibration for the exacerbation module

We performed calibration for asthma exacerbation to match the rate of asthma-related hospitalizations, which are equivalent to very severe exacerbations, observed in Canada by year, sex, and age (Lee et al., 2022a). Of note, the denominator of the observed rate is the general population, not the asthma population. For each year, sex, and age, we computed the predicted proportion of time spent in each of the asthma control levels and then calculated the predicted annual rate of exacerbations. Under the simplifying assumption that individuals labeled as asthmatic had no past history of very severe exacerbations, we calculated the predicted rate of very severe exacerbation per unit of general population. Subsequently, for each year, sex, and age, we compared it with the observed value to obtain a calibration multiplier value and the logarithm of the multiplier was added to the exacerbation equation as β_0_. Of note, our assumption implies that calibration will result in slightly higher rates than observed.

### Payoffs module

The payoffs module is responsible for assigning utilities and costs. The utility values were measured in the quality-adjusted life year (ranging from 0 [death] to 1 [perfectly healthy]) using EQ-5D (Balestroni and Bertolotti, 2012). We obtained the baseline population average utility values for the Canadian population from a recent study (Yan et al., 2024). As this study does not provide the utility values for less than 18 years of age, we made an assumption that the utility values started at 1 at birth and linearly interpolated to 18 years of age for each sex (where the utilities were 0.881 for females and 0.875 for males).

We modeled disutility values due to having asthma for each control level (Einarson et al., 2015) and due to having exacerbations for each severity level (Aldington and Beasley, 2007; Yaghoubi et al., 2020). Disutility of well-controlled asthma was 0.06. This value was 0.09 for partially-controlled asthma and 0.10 for uncontrolled asthma. Disutility of having a mild exacerbation for a year was 0.32. This value was 0.44 for moderate exacerbation and 0.56 for very severe exacerbation. To estimate disutility for severe exacerbation, we took the midpoint between the disutility values of moderate and very severe exacerbations: 0.50. We assumed a mild exacerbation lasted one week (leading to disutility of 0.32 * 1/52 = 0.006) and exacerbations of other severity levels lasted two weeks (Aldington and Beasley, 2007).

Direct annual costs due to having asthma by the control levels and having exacerbation by the severity levels were extracted from Yaghoubi et al. (2020). The costs were converted from 2018 USD to 2023 CAD using historical inflation (1 USD in 2018 = 1.22 USD in 2023; 1 USD = 1.36 CAD in 2023). To estimate the direct cost of having a severe exacerbation, we took the exponential of the average of the log costs of moderate and very severe exacerbations.

### Microsimulation: pseudocode

We have described how the structural equations in each of the five main modules were constructed and used to simulate individual-level characteristics and asthma events in the microsimulation. We explain in more detail how these modules are utilized through pseudocode of the core of the microsimulation (Figure 7).

**Figure 7:**
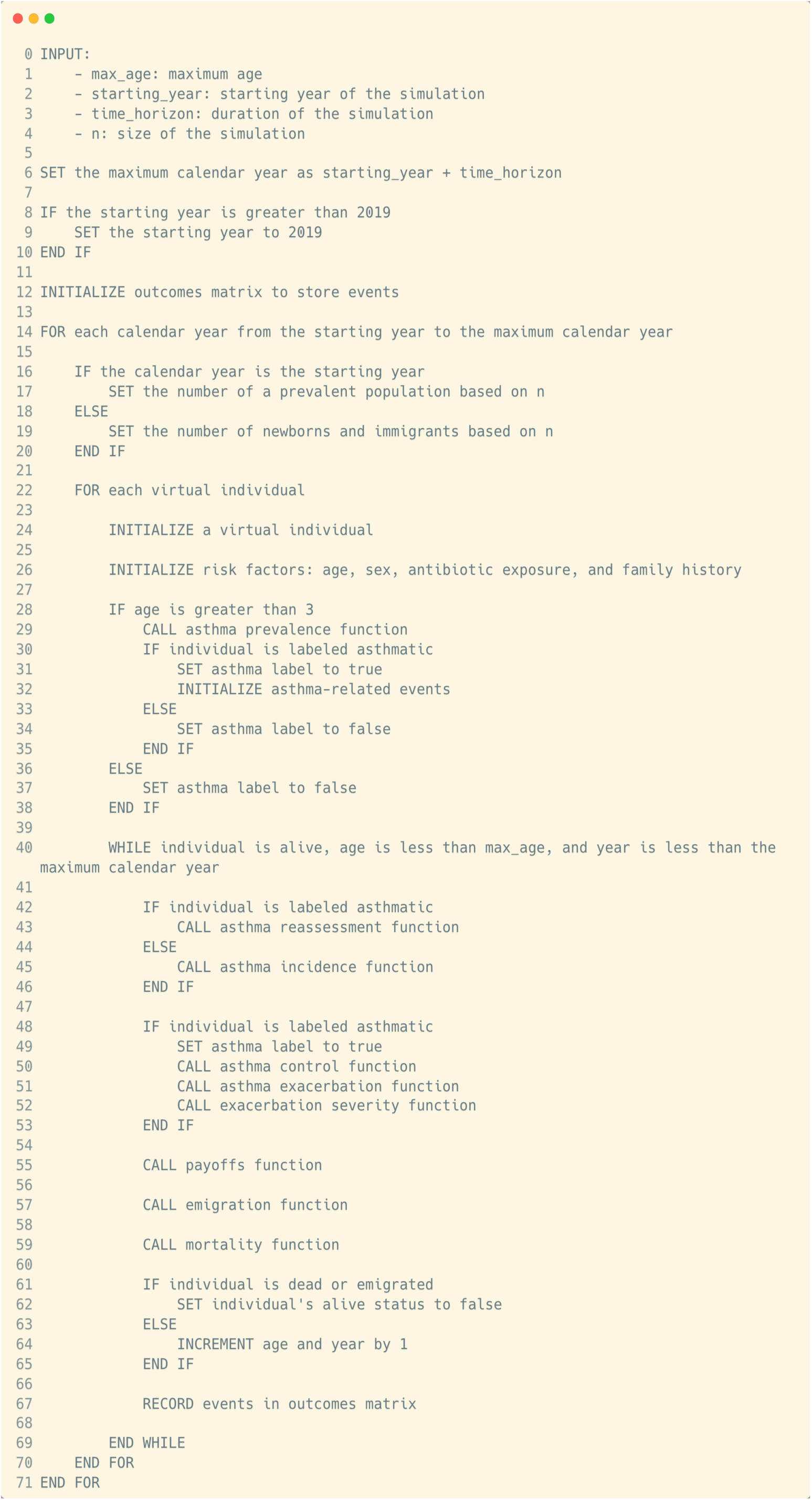
Pseudocode of the asthma model.

The input parameters of the microsimulation (lines 0–4) determine the maximum age of each virtual individual examined (110 is the default), the starting year of the simulation, the period for which the simulation is run, and the size of the population (e.g., 10% of the whole population). Subsequently, the maximum calendar year is calculated (line 6), and the starting year is set to 2019 if it exceeds 2019 (lines 8–10) as explained in the demographics module. We also initialize an array to store events over all virtual individuals.

For each calendar year, we determine how many virtual individuals to simulate based on the demographics module (lines 16–20). If it is the starting year, we need to simulate the entire initial population. If not, we simply need to simulate newborns and immigrants.

Then we simulate each virtual individual (line 22) and begin with initializing their risk factors (line 26). Sex and age are assigned based on the demographics module, and family history of asthma at birth and infant antibiotic exposure are simulated based on the risk factors module. Next, we determine their asthma attributes from the previous year (lines 28–38). This happens only if their age is greater than 3 years since asthma attributes are not given for individuals less than 3 years of age. We assign the asthma label based on the asthma prevalence equation and simulate asthma control and exacerbations if they are labeled as asthmatic. This is the end of the initialization of the demographic and disease characteristics for a virtual individual.

Now the virtual individual enters a conditional loop (line 40). They exit the loop if they meet any of the conditions: they are dead or emigrate, their age exceeds the prespecified maximum age, or their current calendar year is greater than the prespecified maximum year. In each time cycle, we first run the asthma occurrence module to determine whether they are labeled as asthmatic (lines 42–46). If they are labeled as asthmatic, then we simulate asthma-related outcomes, namely asthma control and exacerbations (lines 48– 53). Then we call the payoffs module to evaluate their utility and costs (line 55). We check whether they emigrate via the emigration submodule (line 57) or die via the mortality submodule (line 59). If they die or emigrate, we update their status so they will exit the loop (line 62). Otherwise, we increment their age and year by 1 (line 64).

After all the virtual individuals are simulated and exit the simulation across all the years, we stop the simulation and examine the outcomes matrix.

### Implementation

Julia was chosen as the main engine for running the model. Julia offers the flexibility of an interpretative language (e.g., R and Python) and the near-efficiency of a low-level compiled language (e.g., C++). This has been a major design feature of Julia, making it a powerful tool for computationally intensive calculations without using a low-level language (Lee and Sadatsafavi, 2021). Despite these attractive features, Julia does not yet have the same popularity of Python and R. In particular, open-source modeling in R is receiving attention in the health policymaking community. As such, to facilitate the uptake of the model, we also provided instructions on how to use the model in R. The model is publicly available on the GitHub repository (https://github.com/tyhlee/LEAP.jl).

## Results

### Demographics module

The adjustment β_5,6_ for mortality in (1) was 0.0150 for males and 0.0135 for females. The asthma model simulated mortality robustly as illustrated by close alignment of the simulated and estimated (prior to 2020) or projected (2020 onwards) values from Statistics Canada (Figure 8). After incorporating the adjustment in mortality and net immigrants and emigrants, the model also generated the population growth and aging as estimated or predicted by Statistics Canada (Figure 9).

**Figure 8:**
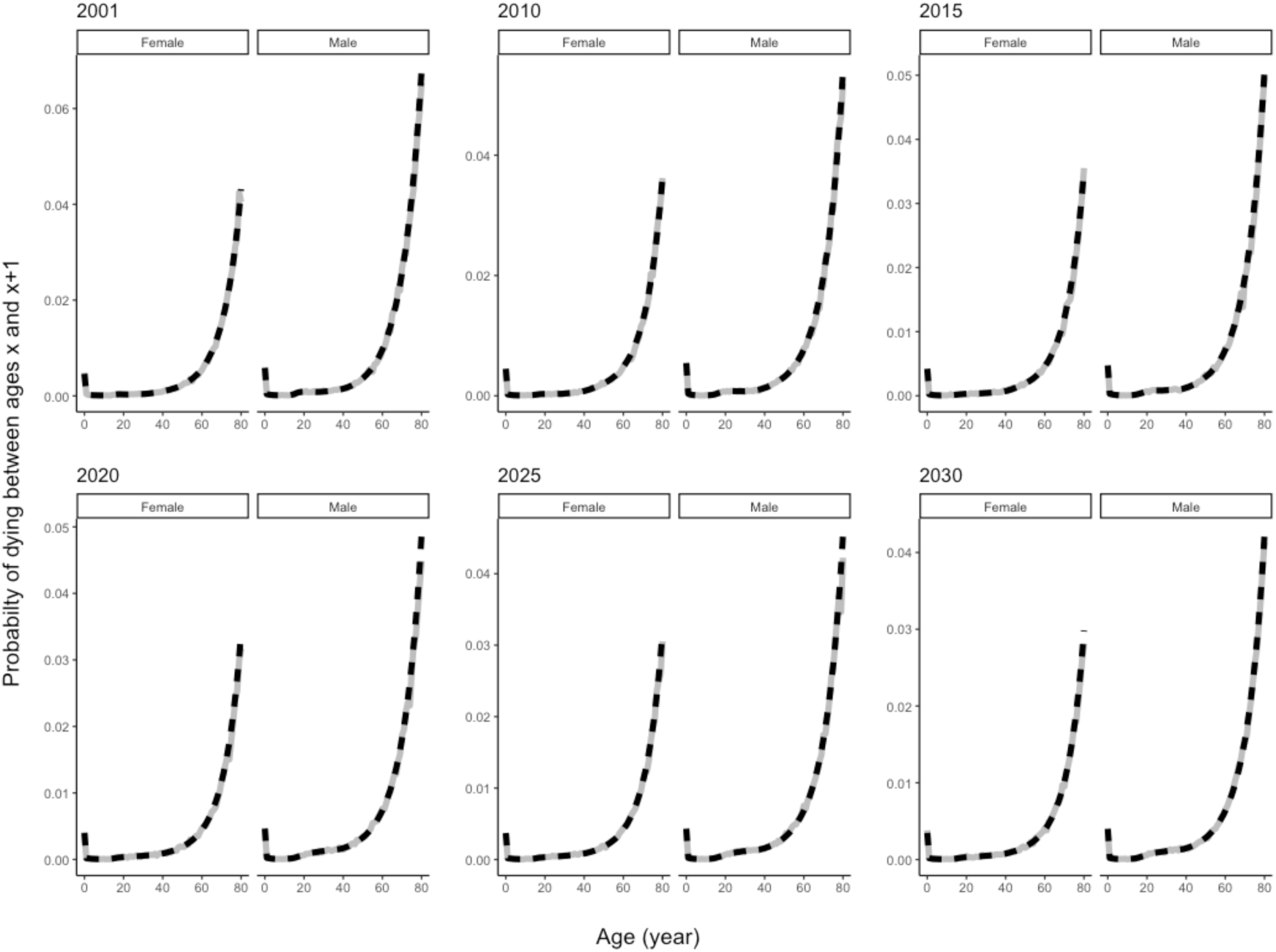
Mortality by sex (left: males; right: females) for the model (grey solid) and Statistics Canada (black dashed).

**Figure 9:**
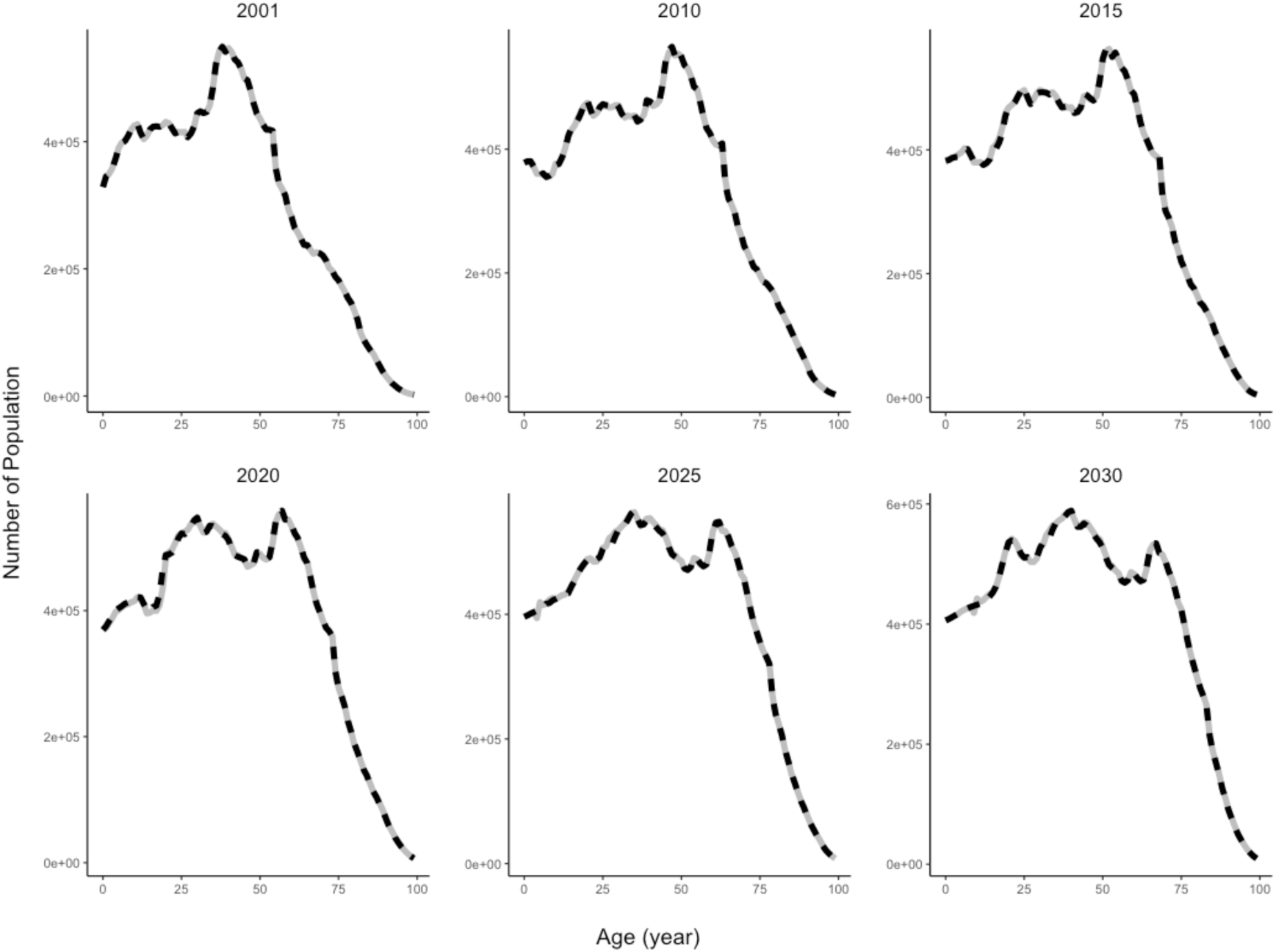
Population by age across selected years from the model (grey solid) and from Statistics Canada (black dashed).

### Risk factors module

The asthma model correctly produced the distribution of risk factors. The mean proportion of simulated individuals with family history of asthma was 29.3% (95% CI: 29.2,29.3) with the true value of 29.3%. The simulated trends in antibiotic exposure in the first year of life followed the observed trends well, and plateaued at 50 per 1,000 as programmed (Figure 10).

**Figure 10:**
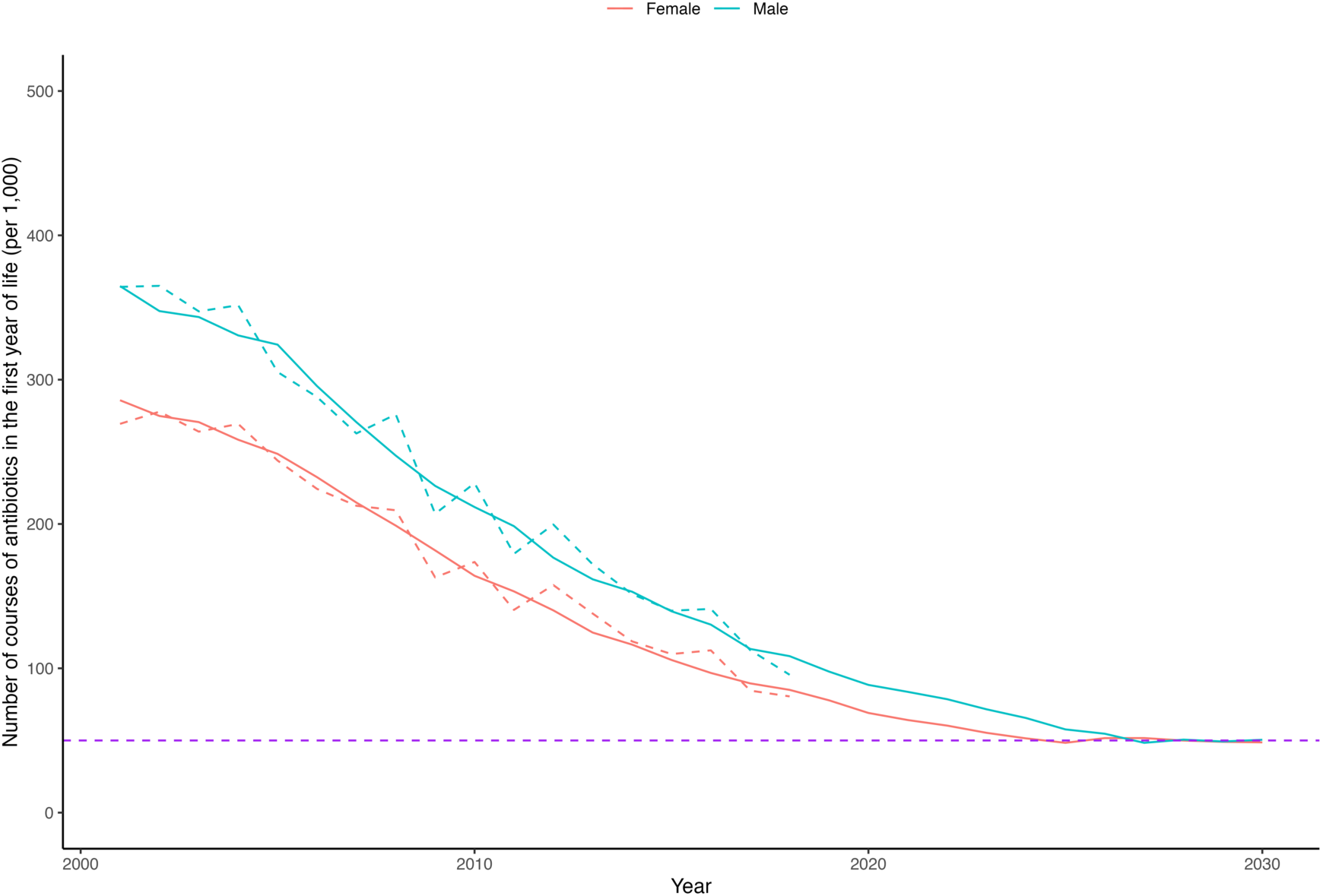
Rate of antibiotic prescriptions by sex (red: females; blue: males) for simulated (solid) and target values (dotted), with the floor rate of 50 per 1,000 (purple).

### Asthma occurrence module

After incorporating calibration, the asthma model produced asthma prevalence rates in close alignment with target values (estimated or projected) prior to the stabilization year, 2025 (Figure 11). After the stabilization year, we noticed the asthma model started deviating slightly from the target values. The reason was that the asthma reassessment probability hit the upper bound of 1 among the young adult population (when solving (1) led to a value slightly greater than 1, e.g., 1.04). This occurred when the asthma prevalence was much higher than the previous year and the asthma incidence was not high enough to match the desired number of individuals labeled as asthmatic. As such, the model generated fewer individuals labeled as asthmatic after the stabilization year. Consequently, the model would be reliable until 2030 or so. On the other hand, excluding the reference levels (i.e., no family history of asthma at birth and no exposure to antibiotics in the first year of life), we found that differences between the target and simulated log ORs for asthma prevalence were just slightly larger than expected. With the ideal value being 0, the median and mean differences were 0.05 and 0.07, respectively, implying simulated values on average were modestly lower than the target values. The extreme differences (the minimum and maximum values are –1.40 and 1.72, respectively) below the lower 1% quantile (–0.48) and above the upper 99% quantile (0.93) all occurred in combinations of year, sex, age, and risk factors with sparse data (i.e., ORs for 3 or more courses of antibiotics in the first year of life).

**Figure 11:**
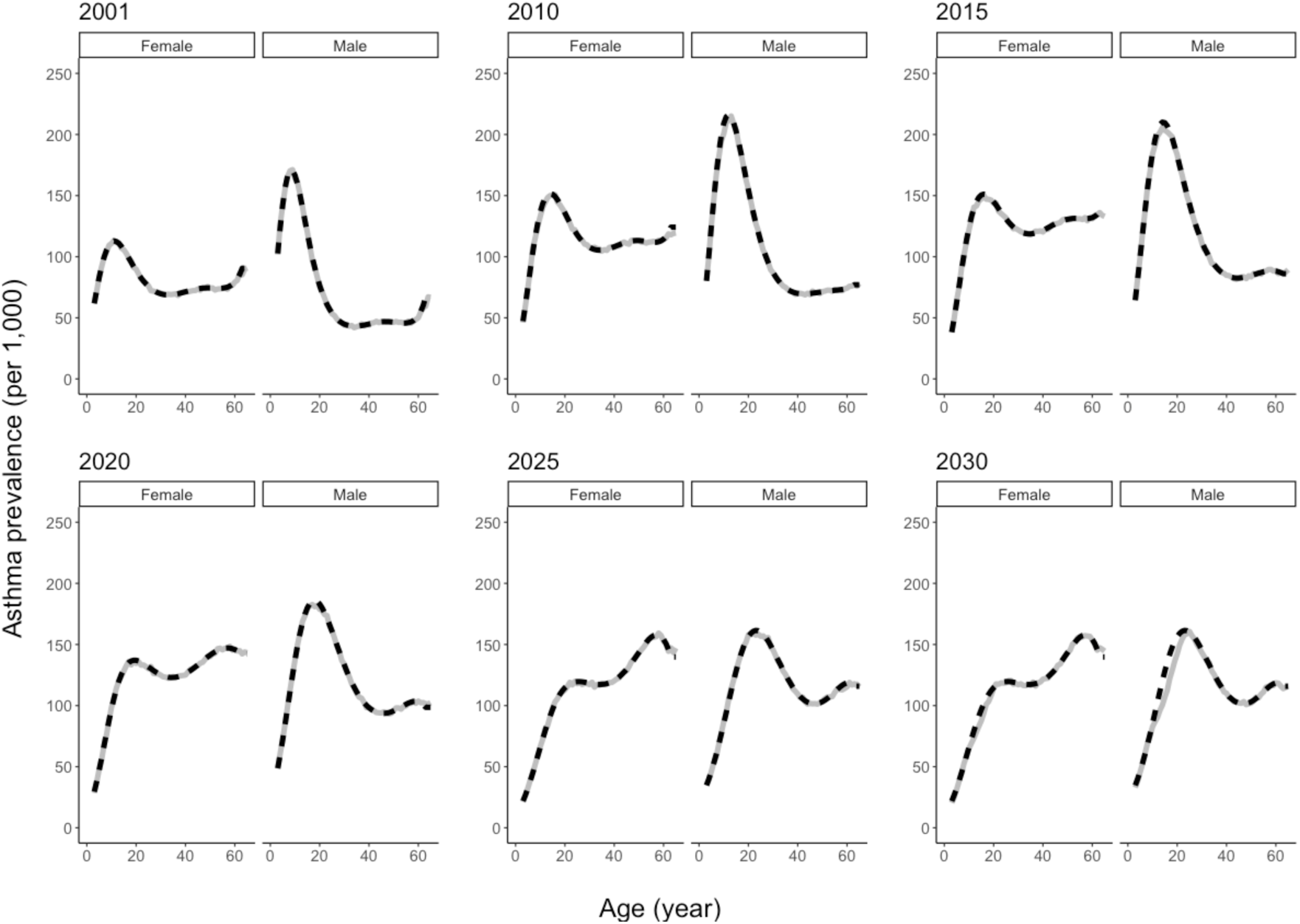
Asthma prevalence rates per 1,000 general population by sex (left: females; right: males) from the model (grey solid) and estimated or projected (black dashed).

### Asthma outcomes module

The asthma model correctly produced the asthma control levels (Figure 12). Asthma severity levels appeared to be correctly modeled (Figure 13); the simulated value of very severe exacerbations should be slightly higher than the target value, as the risk of very severe exacerbations was increased for individuals with past history of very severe exacerbation. Consequently, the asthma model produced slightly higher annual rates of very severe exacerbations than observed values (Figure 14). However, differences in the ratio of target and simulated very severe exacerbation rates on the log scale were modest, ranging from –0.19 to 0.07 with median and mean values of –0.08 and –0.09.

**Figure 12:**
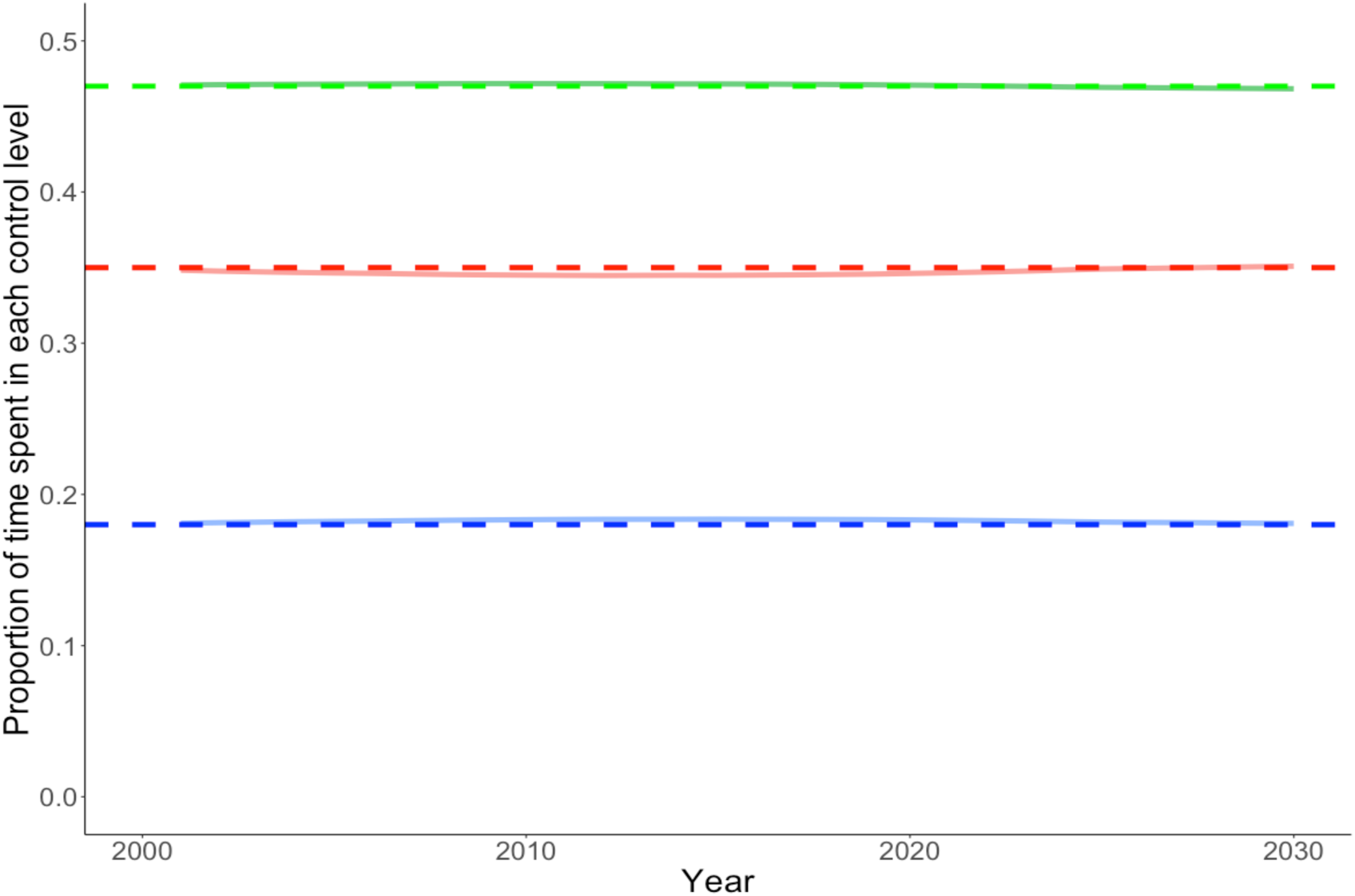
Asthma control levels (red: well-controlled; green: partially controlled; blue: uncontrolled) by the model (solid) and target (dashed).

**Figure 13:**
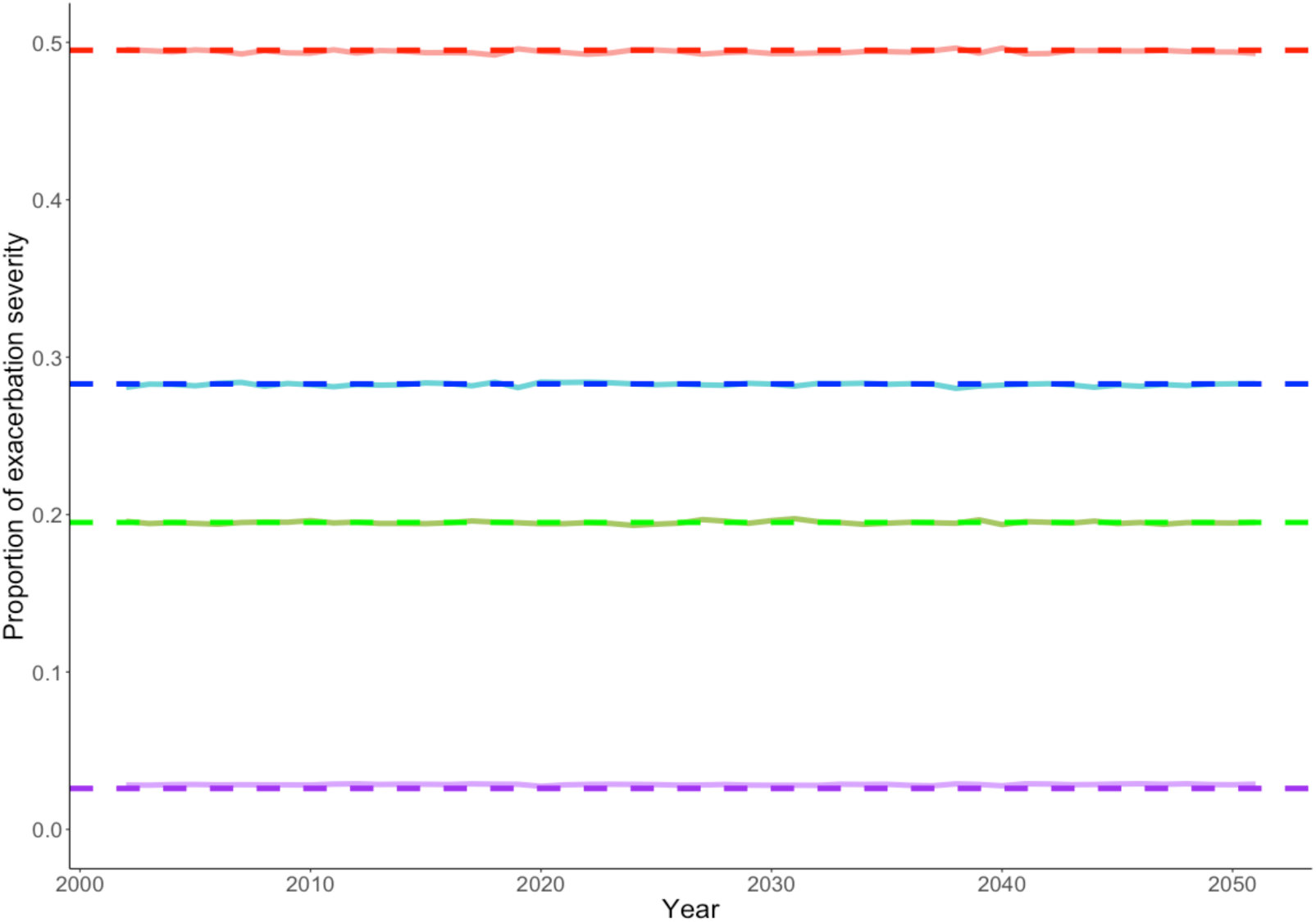
Simulated (solid) and target (dotted) asthma exacerbation severity levels (red: mild; blue: moderate; green: severe; purple: very severe).

**Figure 14:**
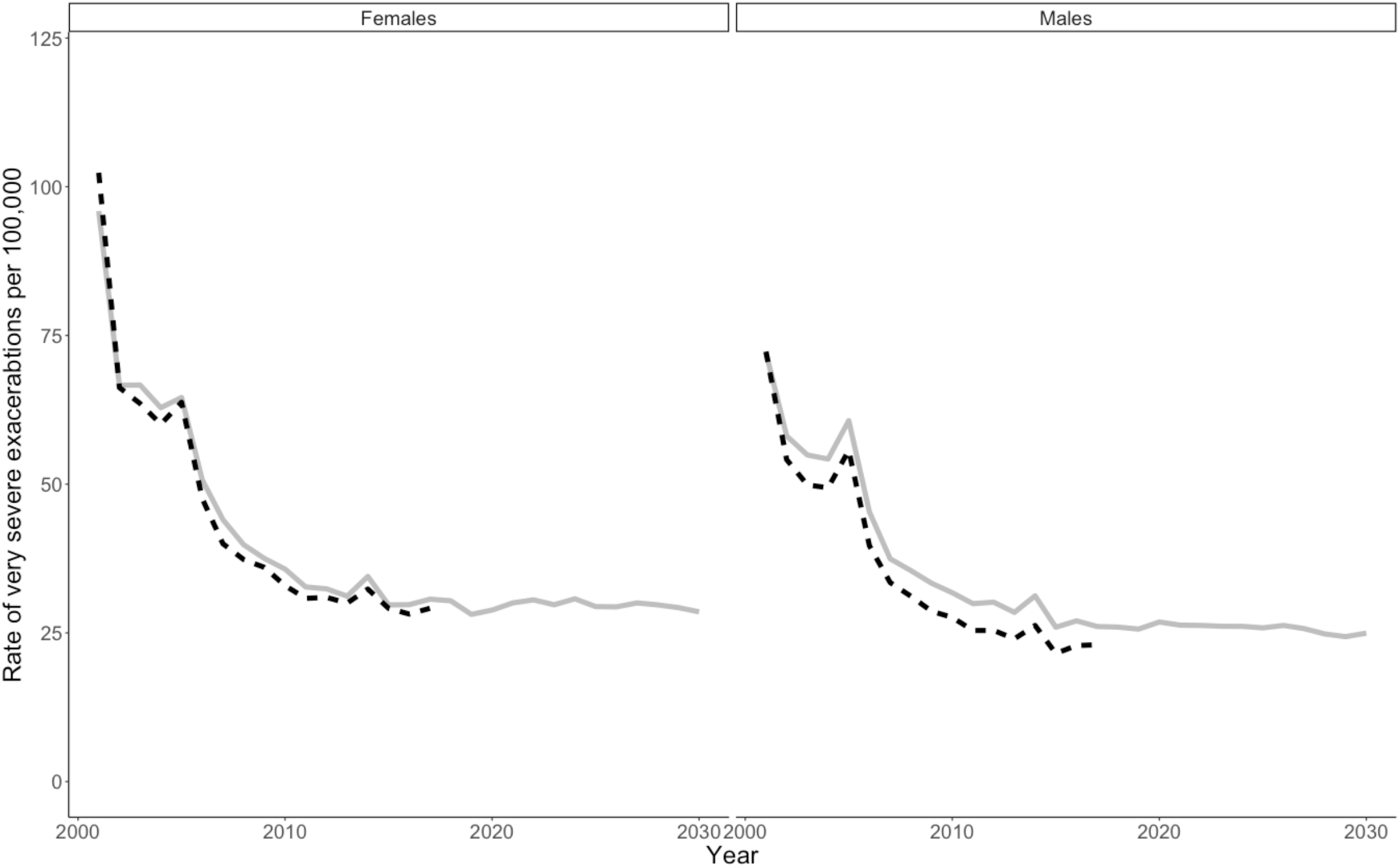
Comparison of simulated (grey solid) and target (black dotted) very severe asthma exacerbation (asthma-related hospital admissions) per 100,000 general population by sex (left: females; right: males) across years.

## Discussion

We developed the first policy model for asthma in Canada with a focus on early childhood asthma and prevention. As recommended by the steering committee, the model was designed to provide a unified framework under which different interventions can be consistently and concurrently compared. More specifically, the following several design choices were made. First, we used an open-population structure that enables modeling of realistic aspects of population aging and adopting health interventions, such as market penetration. Further, we used an individual-level simulation platform to robustly model the vast parameter space created by the many combinations of patient and disease characteristics and event histories. To populate this model, we used multiple data sources and evidence synthesis methods to obtain the highest quality of evidence and generalizability. This included a secondary analysis of several population-based and clinical datasets to estimate disease prevalence, variations in control levels, and exacerbation rate and severity.

Major focus was placed on designing an easily accessible, transparent, and expandable platform. The asthma model was built in a modular approach, with each module composed of submodules in a hierarchical fashion. The modular design offers great flexibility and reusability. Since each module is developed separately, it does not require knowledge of other modules for further development. In addition, technical bugs are easier to detect and thus repair. Code refactoring is also easier in the long run with the modular approach. However, it comes with the cost of increased programming complexity and the need for rigorous documentation.

We found that the asthma model performed satisfactorily in the face and internal validation studies. With calibrated mortality and net immigrants and emigrants, the model replicated the population growth quite well. The marginal asthma prevalence was replicated well up to the stabilization year, but was underestimated among young adults after the stabilization due to the limitation of the framework with asthma reassessment. One could overcome this limitation by modeling true asthma states and introducing an asthma diagnosis module as a means to calibration (i.e., in addition to reassessment and incidence, misdiagnosis can be used to generate asthmatic individuals). Calibration of asthma prevalence by the risk factors was satisfactory in general although the assumption that the risk factor equations between asthma prevalence and incidence are equal at age of 3 years might be unnecessarily strong. For better calibration, one could relax this assumption and allow a more flexible form of the risk factor equations for asthma incidence (e.g., optimize β_0_’s *f_inc_* and *g_inc_*, and include interaction effects among the risk factors). As for the asthma outcomes module, asthma control and severity levels were replicated well. The simulated rate of very severe exacerbations was slightly higher than observed values. A better calibration method that more fully accounts for the past history of very severe exacerbations would eliminate this deviation.

The limitations of this work should be recognized. Currently the natural disease course of asthma is modeled after labeled, rather than true, asthma states. True asthma states should be modeled in the future upon accumulation of sufficient evidence on true asthma states (either through expert knowledge elicitation and/or analysis of data on true asthma states), which would enable evaluation of interventions regarding asthma diagnosis. We have currently incorporated a selective subset of asthma risk factors, but many more risk factors, such as exposure to air pollution, need to be developed to comprehensively capture the disease course of asthma. Some important modules also need to be developed. Particularly important is an asthma management module that can model various asthma management and treatment strategies, and their realistic aspects (e.g., suboptimal adherence). Of note, currently, treatment is ‘implicit’ in the model as the asthma outcomes module is based on data sources that included patients under current standard of care. This approach was adopted as the first scenarios for the use of this model pertain to asthma prevention policies. However, comparative analysis of the outcomes under different asthma management strategies will require creating an explicit treatment module. Immigration and emigration were modeled as a means to calibrate for the population growth and projection. This implies policies surrounding immigration and emigration should not be evaluated with the current version of the model until further development. Further, interactions among virtual individuals were not modeled. This means the model cannot evaluate interventions that target family units (such as subsidizing air filters for families with multiple asthma patients). More importantly, the current version of the asthma model does not account for uncertainty in parameters (Thom, 2022). While this is a current limitation, it was a conscious choice to initially emphasize proper modeling of the structure. Future work requires specification of the parameter distributions and updating calibration procedures that are specifically tuned for probabilistic projections (Shewmaker et al., 2022).

Development of the reference policy model is one step towards rigorous evaluation of interventions for asthma. It addresses the concerns with existing asthma models on generalizability, transparency, accessibility, and granularity in details (Ehteshami-Afshar et al., 2019). It represents consolidated efforts in evidence generation, evidence synthesis, and validation. With further development and validation, the value of the asthma reference policy model will continue to be enhanced and will present an invaluable platform on which resources can be focused to inform optimal investment in asthma prevention and care.

## Data Availability

All data open to the public are available online at https://github.com/tyhlee/LEAP.jl.
The CHILD study cohort data is not open to the public. One can obtain access by following the procedures: https://childstudy.ca/for-researchers/data-access/.

## Appendix

### 1. Best population scenario

*Statistics Canada provides nine scenarios for the demographics projection starting in 2020. The medium growth 3 (M3) scenario resulted in the lowest root mean squared error of the total population at the national level when compared with the observed data in 2020 and 2021*.

| Population projection scenario | Root mean squared error |
| --- | --- |
| Medium growth 3 | 71708 |
| Medium growth 2 | 71714 |
| Medium growth 1 | 71714 |
| Medium growth 4 | 71714 |
| Medium growth 5 | 71741 |
| Slow aging | 102175 |
| High growth | 122869 |
| Fast aging | 132307 |
| Low growth | 152424 |

### 2. Life expectancy calculation

Given the probability of death for each sex and age in 2020 and a value for β_5,6_, the probability of death for each sex and age in 2068 given by (1) allows evaluation of the corresponding projected life expectancy at birth (our target values are 87.0 years for males and 90.1 years for females). The life expectancy in year *t* is calculated based on the current life table for year *t*, following the notations and definitions in Strauss et al. (2023). Let *I*(*x*) be the number of persons alive at age *x*, *d*(*x*) = *I*(*x*) − *I*(*x* + 1) be the number of deaths in the interval (*x*, *x* + 1) for persons alive at age *x*, *q*(*x*) be the probability of dying at age *x*, *L*(*x*) be the total number of person-years lived by the cohort from age *x* to *x* + 1, and *T*(*x*) be the total number of person-years lived by the cohort from age *x* until all members of the cohort have died (i.e., the sum of *L*(*x*) from age *x* to the maximum age, which is 110 years in the Canadian life table generated by Statistics Canada). Then *e*(*x*), the remaining life expectancy of persons alive at age *x*, is calculated as *e*(*x*) = *T*(*x*)/*I*(*x*).

Life expectancy at birth is *e*(0) = *T*(0)/*I*(0). To calculate *T*(0), *L*(*x*) is needed for all ages *x*. Note that *L*(*x*) is the sum of the years lived by the *I*(*x* + 1) persons who survive the interval, and the *d*(*x*) persons who died during the interval. The former contribute exactly 1 year each, while the latter contribute, on average, approximately half a year. For the boundary of ages of 0 and 110, a smaller contribution (< 0.5) is usually used for the former, and a larger contribution (between 1 and 2) is usually used to for the latter (since they live longer than 110 years but not very much longer). In general, with *w*(*x*) denoting the contribution made by members of the cohort who die at age *x*, *L*(*x*) = *I*(*x* + 1) + *w*(*x*)*d*(*x*). We chose, by trial and error, *w*(*x*) = 0.2 for *x* = 0, 1.4 for *x* = 110, and 0.5 for all other values of *x*. This choice of *w*(*x*) matched the calculation of the life expectancy at birth for the 2020 life table by Statistics Canada.

In summary, using (1) and initial values of β*_sex_*, we first calculated *q*(*x*) for all ages *x* in 2068. We used *I*(0) = 100,00 (other values can be used as the value of *I*(0) cancels out during evaluation of life expectancy) and calculated *d*(*x*) = *I*(*x*)*q*(*x*) and *L*(*x*) = *I*(*x* + 1) + *w*(*x*)*d*(*x*). Then we computed *T*(*x*) and finally *e*(*x*). Using the uniroot solver, we found the optimal values β*_sex_* that matched our target life expectancy values at birth in 2068.

### 3. Calibration for asthma incidence

Calibration for the asthma incidence equation is difficult since the effects of risk factors are unknown and need to be estimated such that both the marginal asthma incidence and the odds ratios in the asthma prevalence equation remain unchanged. The former can be achieved with the method used for the asthma prevalence calibration, but the latter involves several steps. As described in the main text, our goal is to optimize β_1’_’s in *f_inc_* and *g_prev_*. Required inputs are the odds ratios for asthma prevalence, calibrated prevalence, and the probability distribution of the risk factor(s) from the previous time step, and odds ratios for asthma prevalence, crude incidence, and reassessment probabilities from the current time step. We also need to specify initial values for β_1_’s (we used the corresponding values from *f_prev_* and *g_prev_*). The first step is to compute, for each combination of sex and age, an odds ratio for being labeled asthmatic for each level *x* of the risk factor relative to its reference level for the current time step. Put differently, we need a 2×2 contingency table of proportions, not counts, for that level *x*.

To construct this contingency table, we first need the contingency table of level *x* from the previous time step. Given an odds ratio of level *x*, the proportion of individuals labeled as asthmatic (among individuals in level *x* and the reference level), and the proportion of individuals being at level *x* from the previous time step, the table can be obtained using a method by Bonett (2007). Let *a*_0_, *b*_0_, *c*_0_, *d*_0_ represent this table, with *a*_0_ and *c*_0_ being the proportion of individuals not labeled as asthmatic at the reference level and at level *x*, and *b*_0_and *d*_0_being the proportion of individuals labeled as asthmatic at the reference level and at level *x*.

As described in the main text, specifying initial values for the ORs in the incidence equation followed by calibration for the overall incidence rate provides a revised incidence equation. With the incidence rate for the reference level (*inc*_0_) and level *x* (*inc_x_*), we now construct the corresponding contingency table for the current time step. The first cell *a* is equal to the proportion of individuals at the reference level who either lose their asthma label (*b*_0_1 − *p*(*reassessment*))) or are not newly labeled as asthmatic (*a*_0_(1 – *inc*_0_)). Having calculated *a*, the second cell *b* can be obtained by subtraction: *b* = *a*_0_ + *b*_0_ − *a*. The cells *c* and *d* are similarly obtained from *c*_0_, *d*_0_, and *inc_x_*. We then compute the odds ratio, compare it to the target odds ratio for level *x*, record the absolute difference. To find a set of values of β_1’_’s in the asthma incidence equation, we aggregated the absolute differences over all combinations of year (up to the stabilizing year, 2025), sex, and age, and used the iterative BFGS algorithm until the aggregated difference was less than 10^-10^.

## Notes

### Competing Interest Statement

The authors have declared no competing interest.

### Author Declarations

This study was approved by the institutional review board of the University of British Columbia, Vancouver (H22-00571).

